# Plasma p217+tau vs NAV4694 amyloid and MK6240 tau PET across the Alzheimer continuum

**DOI:** 10.1101/2022.02.16.22271024

**Authors:** Vincent Doré, James D. Doecke, Ziad S. Saad, Gallen Triana-Baltzer, Randy Slemmon, Natasha Krishnadas, Pierrick Bourgeat, Kun Huang, Samantha Burnham, Christopher Fowler, Stephanie R. Rainey-Smith, Ashley Bush, Larry Ward, Jo Robertson, Ralph N. Martins, Colin L. Masters, Victor L. Villemagne, Jurgen Fripp, Hartmuth C. Kolb, Christopher C. Rowe

**Affiliations:** The Australian e-Health Research Centre, CSIRO, Melbourne, Victoria, Australia; Department of Molecular Imaging & Therapy, Austin Health, Melbourne, Victoria, Australia; The Australian e-Health Research Centre, CSIRO, Brisbane, Queensland, Australia; Neuroscience Biomarkers, Janssen Research and Development, La Jolla, CA, USA; The Florey Institute of Neuroscience and Mental Health, Melbourne, Victoria, Australia; Edith Cowan University, Perth, Australia; Florey Department of Neuroscience and Mental Health, The University of Melbourne, Melbourne, Victoria, Australia; Centre for Healthy Ageing, Health Futures Institute, Murdoch University, Murdoch, WA, Australia; McCusker Alzheimer’s Research Foundation, Nedlands, Perth, Australia; Department of Psychiatry, University of Pittsburgh, Pittsburgh, PA, USA

**Keywords:** Plasma p-tau217, Tau imaging, Aβ-amyloid imaging, Alzheimer’s disease, positron emission tomography, blood biomarkers, blood diagnostic for Alzheimer’s disease, phosphorylated tau, amyloid plaque, paired helical filaments

## Abstract

**INTRODUCTION:** We evaluated a new Simoa plasma assay for phosphorylated tau at aa217 enhanced by additional ptau sites (p217+tau).

**METHODS:** Plasma p217+tau levels were compared to ^18^F-NAV4694 amyloid-beta (Aβ) PET and ^18^F-MK6240 tau PET in 174 cognitively impaired (CI) and 223 cognitively unimpaired (CU) participants.

**RESULTS:** Compared to Aβ-CU, the plasma levels of p217+tau increased two-fold in Aβ+ CU and 3.5-fold in Aβ+ CI. In Aβ-the p217+tau levels did not significantly differ between CU, MCI or dementia. P217+tau correlated with Aβ centiloids ρ=0.67 (CI 0.64; CU 0.45) and tau SUVR_MT_ ρ=0.63 (CI 0.69; CU 0.34). Area under curve (AUC) for AD vs Aβ-CU was 0.94, for AD vs other dementia was 0.93, for Aβ+ vs Aβ– PET was 0.89 and for tau+ vs tau-PET was 0.89.

**DISCUSSION:** Plasma p217+tau levels elevate early in the AD continuum and correlate well with Aβ and tau PET.

**Research in Context:**

1. Systematic review: The authors reviewed the literature using PubMed, meeting abstracts and presentations. Plasma phospho-tau measures compare well to PET and post-mortem across the continuum of AD but accuracy varies across ptau target sites and assay methods. There are no reports comparing PET to plasma assays targeting multiple sites of tau phosphorylation as typically found in AD. The p217+tau assay targets p217 with binding enhanced by phosphorylation at additional sites such as aa212.
2. Interpretation: Plasma p217+tau elevates early and correlates with both Aβ and tau as measured by PET indicating that tau phosphorylation is an early event in AD and occurs with Aβ deposition. Plasma p217+tau measurement should assist both selection for trials and diagnosis of AD.
3. Future directions: Further validation studies, head-to-head comparison to other assays, assessing the influence of co-morbidities and the ability to measure change in brain Aβ and tau levels are required.

## Introduction

Assessing brain levels of Aβ-amyloid (Aβ) plaque and tau aggregation has a central role in the selection of individuals for clinical trials and assessing the efficacy of therapeutic compounds. It may also aid earlier and more accurate diagnosis of Alzheimer’s disease (AD) [1].

Despite the extremely low concentration of Aβ and tau in plasma, progress in detection methodologies has recently allowed reliable measurement of plasma Aβ_42/40_ and phosphorylated tau (P-tau). Measurement of plasma tau phosphorylated at threonine 181 (P-tau181) was the first to demonstrate good accuracy [2-4]. P-tau181 can differentiate AD from cognitively unimpaired (CU) subjects [5] and from other dementias [6]. P-tau181 was also a good predictor of elevated brain Aβ and tau as measured by PET [7]. Subsequently, plasma P-tau217 and P-tau231 have also shown strong association with AD pathology with accuracy comparable to CSF and PET measures [8, 9] with some evidence of superior performance to plasma P-tau181 [10, 11].

More recently a high sensitivity Simoa assay has been developed using a capture antibody (pT3) that was raised against tau in paired helical filaments (PHF) of AD brain [12,13]. The core requirement for this antibody binding is phosphorylation at aa217 (p217) with enhanced binding when other nearby phosphorylated sites are present, predominantly at aa212 [12]. The recognised epitope is thus referred to as “p217+tau” and its measurement in plasma has demonstrated good concordance with CSF markers of AD in a validation cohort of 227 subjects with Area Under the Curve (AUC) of 0.90 vs CSF Aβ42/40 and 0.95 vs CSF P-tau181 [13].

Plasma p217+tau has not been compared to PET measures. ^18^F-NAV4694 and ^18^F-MK6240 are PET tracers for imaging Aβ plaque and PHF 3R/4R tau aggregates respectively. These recent generation tracers have high target to background ratios giving a wide dynamic range that may improve sensitivity for detection of low levels of Aβ and tau [14, 15].

In this study we evaluated the performance of the novel blood-based biomarker p217+tau against latest generation Aβ and tau PET agents in participants of the Australian Imaging, Biomarkers and Lifestyle study of aging (AIBL) and the Australian Dementia Network (ADNeT) trial screening program.

## Methods

### Participants

Three hundred and ninety-seven participants in the AIBL and ADNeT cohorts had both ^18^F-NAV4694 Aβ PET and ^18^F-MK6240 tau PET. The AIBL cohort recruitment and evaluation is detailed elsewhere [16]. The ADNeT cohort all had mild cognitive impairment (MCI) or mild dementia and were evaluated as per AIBL. A multi-disciplinary panel, blind to all imaging and blood results, classified all participants as cognitively unimpaired (CU) or cognitively impaired (CI) with MCI, Alzheimer’s disease (AD) or non-AD dementia. A diagnosis of CU required performance within 1.5 standard deviations of the published norms for their age group on neuropsychological assessment. A diagnosis of MCI [17] or dementia [18] were assigned according to internationally agreed criteria. For this study, only individuals with a clinical diagnosis of AD and a positive Aβ PET scan were classified as AD. Those with dementia but a negative Aβ scan were classified as non-AD dementia. Institutional ethical review committees approved the AIBL and ADNeT studies and written informed consent was obtained from all participants.

### Image Acquisition

A 20-minute Aβ PET acquisition was conducted 50 minutes post-injection of 200 MBq of ^18^F-NAV4694. On a separate day a 20-minute Tau PET acquisition was conducted 90 minutes post-injection of 185 MBq of ^18^F-MK6240.

Aβ PET scans were spatially normalized using CapAIBL [19] and the standard Centiloid (CL) method was applied [20, 21]. A CL value of 25 was selected to determine a positive Aβ scan (Aβ+) [22-24]. ^18^F-MK6240 tau PET scans were spatially normalized using a CapAIBL PCA-based approach [25] and scaled using the cerebellar cortex as the reference region. ^18^F-MK6240 SUVR values were estimated in two composite regions of interest (ROI); a mesial temporal ROI (Me) comprising entorhinal cortex, hippocampus, parahippocampus and amygdala; and a meta temporal ROI (MT) composed of the Me ROI plus inferior and middle temporal and fusiform gyri. We also estimated ^18^F-MK6240 SUVR in Braak stages IV, V and VI with ROIs derived from the Freesurfer Desikan-Killiany Atlas (see Supplementary Figures 3-4). In the CU group, we additionally estimated ^18^F-MK6240 SUVR in the entorhinal cortex, amygdala, hippocampus and inferior temporal cortex separately. The threshold for tau positivity was the 95^th^ percentile of the Aβ-CU group in each composite ROI and individual mesial temporal regions.

### Plasma Analysis and p217+tau assay design

Fasted blood sampling was performed 3.3±6 months from the time of Aβ PET scan and 1.7±2 months from the tau scan. Plasma from K_2_-EDTA tubes (7.5 mL S-monovette 01.1605.008, Sarstedt) containing pre-added prostaglandin E1 (33 ng/mL of whole blood, Sapphire Biosciences) to prevent platelet activation, a potential source of peripheral amyloid-β, was centrifuged at room temperature at 200g for 10 mins to collect platelet-rich plasma, and then at 800g for 10 mins to provide plasma that was snap frozen within 2 hr of collection was stored in vapour phase liquid nitrogen prior to shipping on dry ice from Australia to Janssen R&D, La Jolla, CA, USA. Plasma ptau217+ assay was performed on a SIngle MOlecular Array (Simoa) HD-X platform blinded to all subject data using the technique previously described [13]. Data analysis was then performed by AIBL investigators.

### Statistical analysis

Demographic and clinical characteristics were compared between Aβ status within clinical classifications using standard Generalised Linear Modelling (GLM) for quantitative features and the Chi-square test for categorical comparisons. Correlation between the plasma p217+tau and CL or tau SUVR was performed using Spearman’s rank-based correlation (Spearman’s Rho [ρ]). Receiver Operating Characteristic AUC analyses were used to explore the discriminative performance of p217+tau. Optimal p217+tau thresholds for detecting Aβ+ and tau+ PET were calculated both using Youden’s Index and two standard deviations above the mean p217+tau levels in Aβ-CU group. Presented are AUC values with 95% confidence intervals (shown in square brackets), sensitivity, and specificity with 95% confidence intervals and positive and negative predictive values (PPV, NPV, not disease prevalence adjusted). Model based performance of p217+tau accounting for age, gender and *APOE* ε4 allele status was done using GLM. AUC values between models were compared using DeLong’s method. Confidence intervals were computed using bootstrap. False discovery rate (FDR) corrected P-values lower than 0.05 were considered significant. Vertex and voxel-based correlations between p217+tau and Aβ and tau images were performed using Spearman’s rank-based correlation.

## Results

### 1. Demographic and Clinical Characteristics

Three participants had a low CL but high Braak stage V/VI tau on ^18^F-MK6240 PET suggesting that, similar to some PiB case reports [26], the Aβ PET were falsely negative. These three subjects all showed clearly elevated plasma p217+tau levels and were kept in the analysis but are displayed with a diamond shape in the scatter plots.

Table 1 presents demographic and clinical characteristic. Of the 397 participants, 56% were cognitively unimpaired (CU), 22% had MCI, and 21% had dementia of which 82% in the dementia group were Aβ+. Overall, 170 (43%) had Aβ+ PET and 143 (36%) had high tau PET in the meta temporal region (T_MT_+). Participants with dementia were approximately 4 years younger than MCI and CU.

**Table 1:** Demographic and clinical characteristics.

|  | **CU**<br>(n=223) | **MCI**<br>(n=91) | **Dementia**<br>(n=83) |
| --- | --- | --- | --- |
| Age | 75.2 (5.7) | 73.6 (8.0) | 70.7 (7.9)<sup>*+</sup> |
| Gender N male (%) | 101 (45.3%) | 51 (56.0%) | 49 (59.0%) |
| Education (years) | 14 (3.0) | 12.5 (3.1)<sup>*</sup> | 12 (3.0)<sup>*</sup> |
| APOE ε4+ (%) | 72 (32.3%) | 46 (50.5%)<sup>*</sup> | 46 (55.4%)<sup>*</sup> |
| MMSE Median (IQR) | 29 (2) | 27 (4)<sup>*</sup> | 23 (4)<sup>*+</sup> |
| CDR SoB Median (IQR) | 0.0 (0.0) | 1.0 (1.5)<sup>*</sup> | 4.0 (1)<sup>*+</sup> |
| AIBL PACC Mean (SD) | -0.4 (0.8) | -2.4 (1.1)<sup>*</sup> | -4.3 (2.4)<sup>*+</sup> |
| Centiloid Mean (SD) | 19.5 (38.1) | 72.8 (66.2)<sup>*</sup> | 93.9 (55.6)<sup>*+</sup> |
| Aβ+ N (%) | 46 (20.6%) | 56 (61.4%)<sup>*</sup> | 68 (81.9%)<sup>*+</sup> |
| MK6240 SUVR<sub>MT</sub> Mean (SD) | 1.02 (0.23) | 1.48 (0.61)<sup>*</sup> | 1.93 (0.77)<sup>*+</sup> |
| Tau<sub>MT</sub>+ N(%) | 31 (13.9%) | 48 (52.7%)<sup>*</sup> | 64 (77.1%)<sup>*+</sup> |
| Adjusted HV (SD) | 2.98 (0.26) | 2.78 (0.40)<sup>*</sup> | 2.57 (0.42)<sup>*+</sup> |
| Plasma p217+tau Mean | 87.8 (67.8) | 183.7 (141.0)<sup>*</sup> | 229.9 (150.7)<sup>*+</sup> |
| (SD) |  |  |  |

*\*p-value<0.05 compared to CU, +p-value<0.05 compared to MCI. AIBL PACC is the AIBL preclinical Alzheimer's cognitive composite; T<sub>MT+</sub> means positive for tau in the tau PET meta temporal ROI; HV is hippocampal volume*

Stratification by Aβ status within each clinical classification is provided in Supplementary Table 1.

### 2. Plasma p217+tau Levels in Clinical Groups by Aβ **PET Status**

Figure 1A shows the concentration of plasma p217+tau in the different clinical groups, subdivided into Aβ- and Aβ+. The mean p217+tau concentration in CU Aβ+ participants (162.7fg/ml (SD: 85.8)) was approximately twice that of the CU Aβ-participants (71.2fg/ml (SD: 46.6), p<10^−18^). Mean p217+tau concentration in MCI Aβ+ (252.0 fg/ml (SD: 141.0)) were not significantly different to those with Aβ+ dementia (AD) (259.3 fg/ml (SD: 147.7)) but both groups showed significantly higher concentration than in both CU Aβ+ participants and Aβ-CU, MCI and dementia participants. Levels of p217+tau in the MCI Aβ-(82.1 fg/ml (SD:55.8)) and the Aβ-non-AD dementia (93.4 fg/ml (SD: 45.6)) were not significantly higher than CU Aβ-participants.

**Figure 1:**
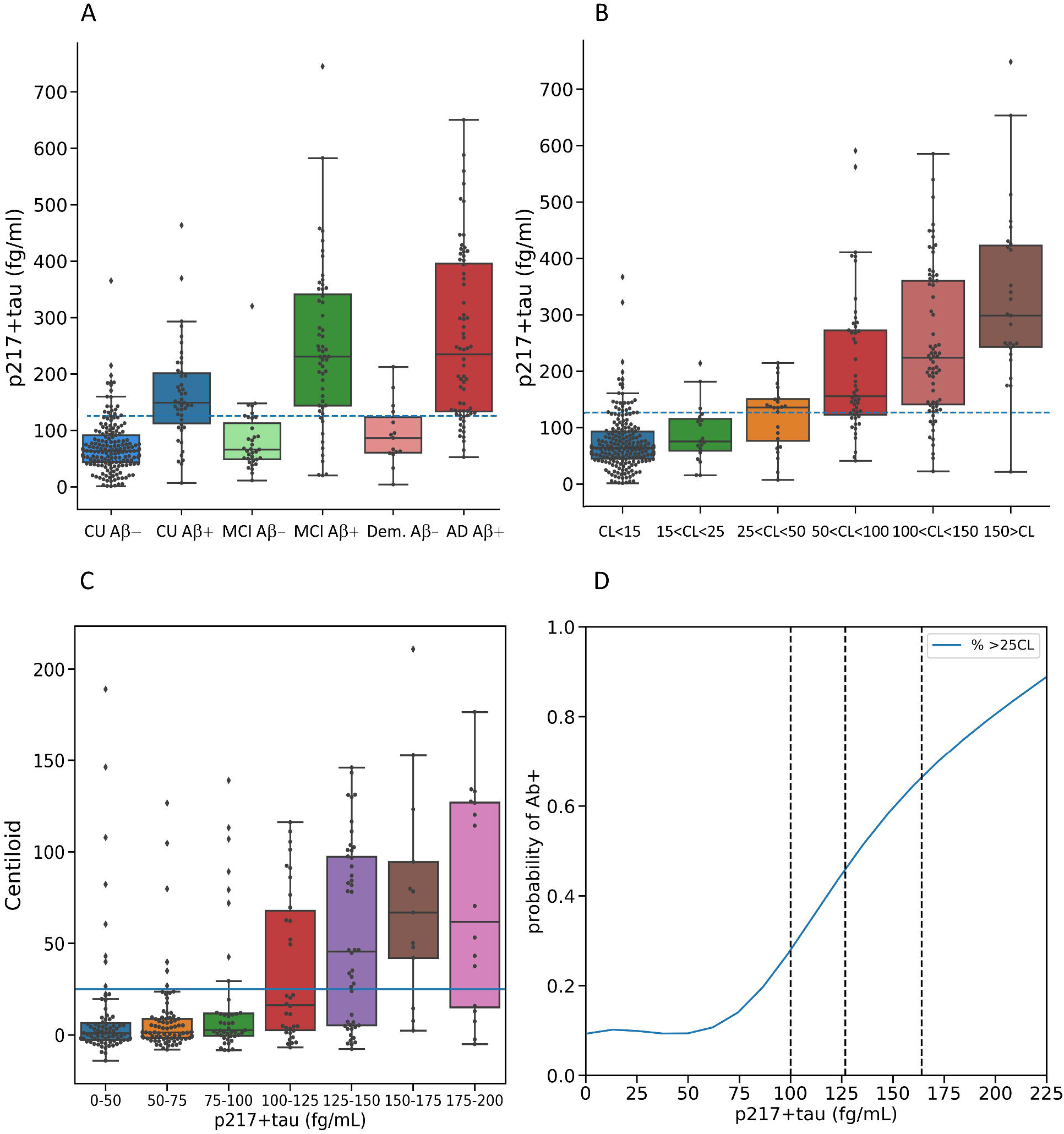
Plasma p217+tau concentrations between A) clinical classification and A□ PET status and B) Centiloid levels of Aβ. The dashed line corresponds to the threshold derived by Youden Index. C) Centiloid results vs p217+tau level in 25 fg/ml intervals, D) probability of being Aβ PET positive vs p217+tau level. Vertical lines in D) are Youden index derived from cognitively unimpaired (CU) (100.3 fg/ml), Youden index derived from total cohort (126.7 fg/ml) and +2.0 SD of the Aβ-ve CU (164 fg/ml).

When subdividing the CL scale into 5 levels (Figure 1B), the ptau217+tau concentration was progressively higher from 25 CL. This progressive increase remained in those who were tau PET negative (see Supplementary Figure 1). Figure 1C shows the Centiloid level with increasing p217+tau level and figure 1D shows the probability of having a positive Aβ scan vs p217+tau level.

### 3. Correlations of plasma p217+tau with Aβ PET and Tau PET

Vertex based analysis (Figure 2 and supplementary figures 6-10) and scatter plots (Figures 3-5) demonstrate moderate to strong correlations between plasma p217+tau and Aβ and tau PET. The correlation in all subjects between p217+tau and Aβ PET CL gave a Spearman’s ρ of 0.67 (p<10^−53^). The correlation was significantly stronger in CI than in CU (CI: ρ=0.64, p<10^−21^; CU: ρ=0.45, p<10^−12^; Z-score difference 2.6) while Figure 2 shows the regional correlation matches the early and predominant locations of Aβ accumulation in AD.

**Figure 2:**
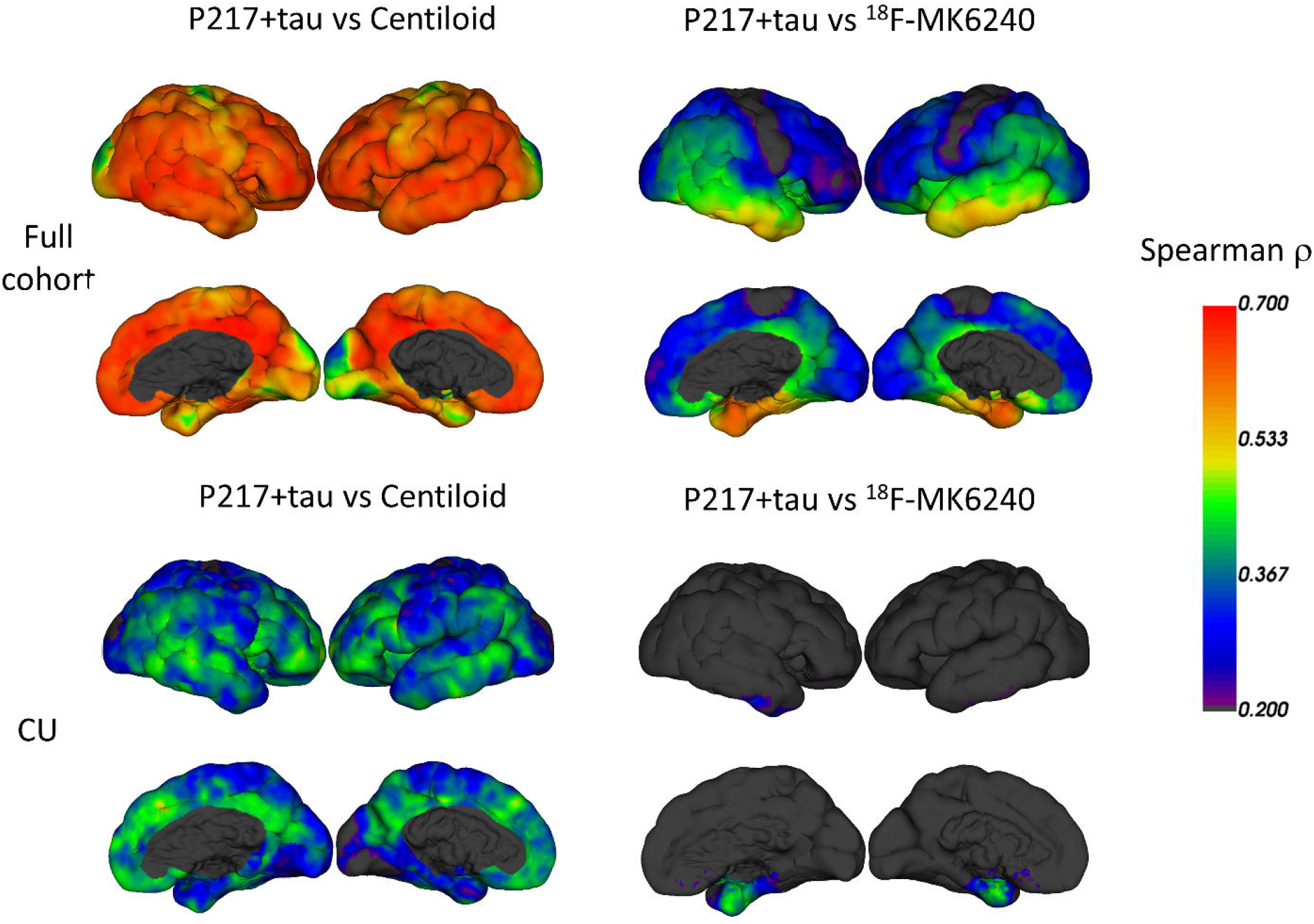
Vertex-based analysis of regional Spearman correlation between plasma p217+tau and Centiloid (left column) and ^18^F-MK6240 SUVR (right column). CU is cognitively unimpaired.

**Figure 3:**
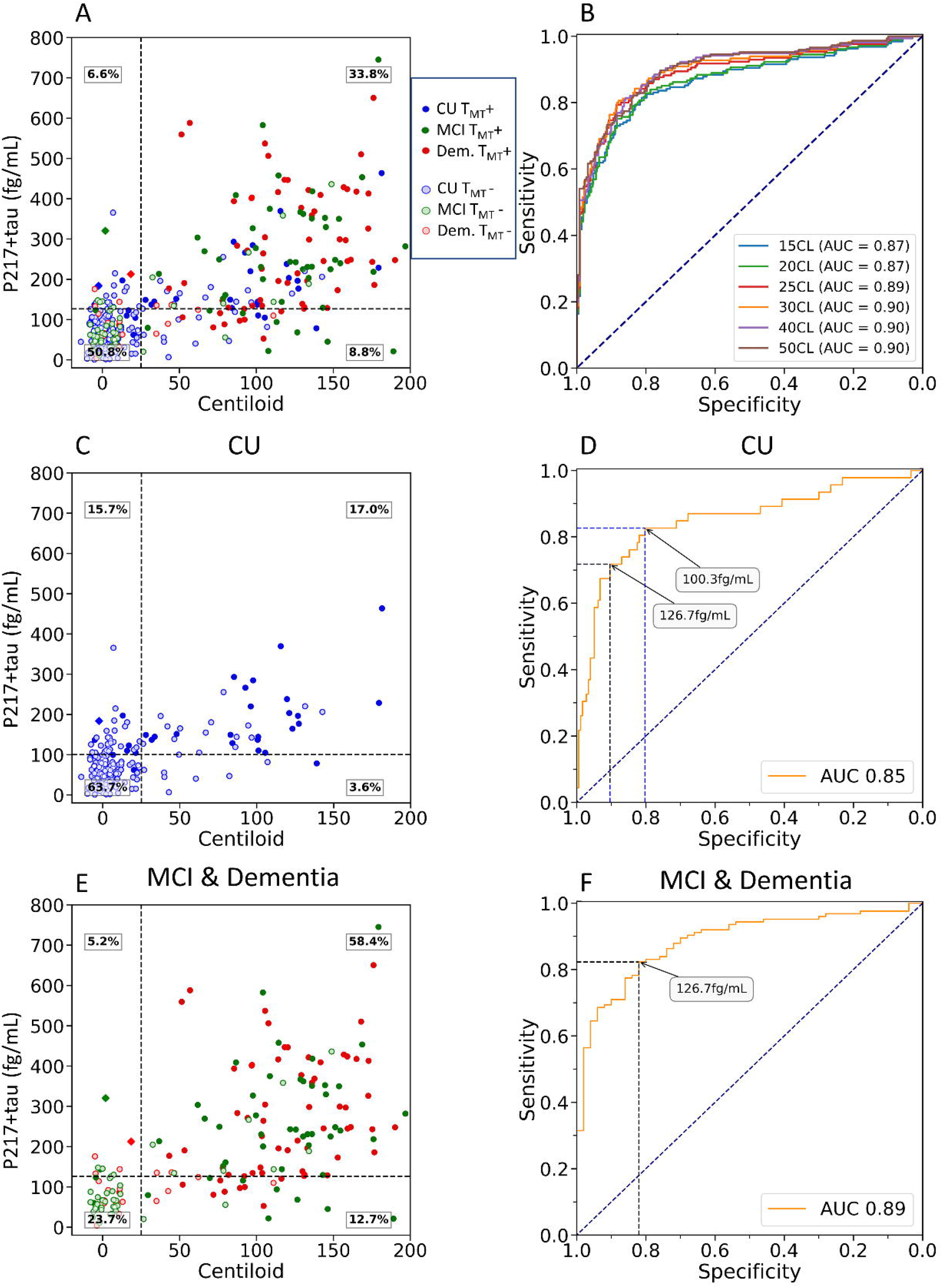
Plasma p217+tau vs Centiloid measure of Aβ. Scatter plot and ROC curve (A&B) with ROC curves using different Centiloid thresholds to define Aβ+ PET; full cohort (A&B), cognitively unimpaired sub-cohort (C&D) and cognitively impaired sub-cohort (E&F). Clinical groups are colour-coded with red for dementia, green for MCI and blue for CU. Solid circles are tau PET positive (T_MT_+). The black dashed horizontal line corresponds to the p217+tau threshold derived from Youden’s index. In C) and D) the CU specific threshold is shown. The CI group threshold was the same as the whole cohort threshold. The diamond shapes are the three Aβ- / T_MT_+ subjects.

**Figure 4:**
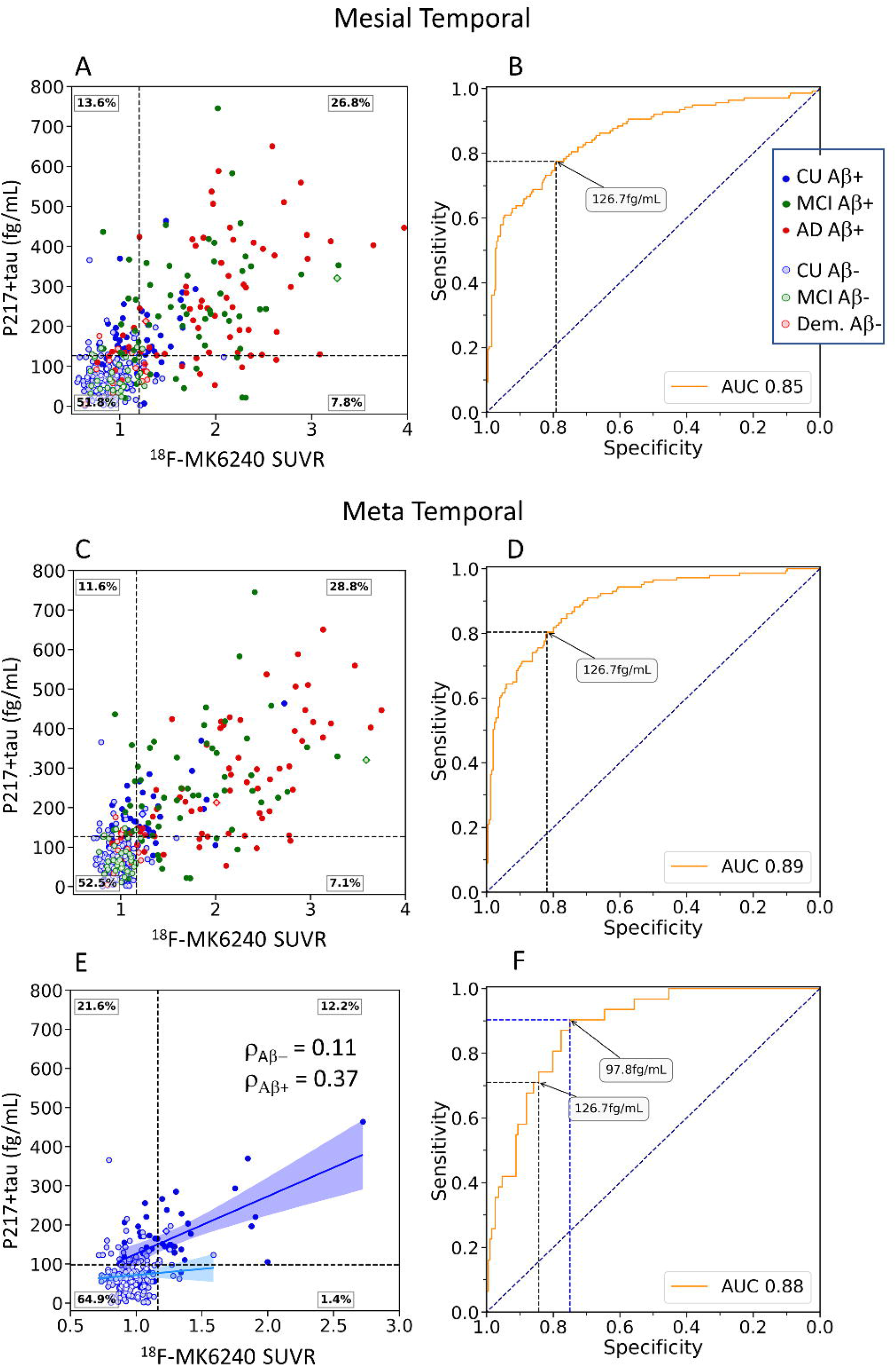
Plasma p217+tau vs PET SUVR measures of tau. Scatter plot and ROC curve for mesial temporal ROI (A&B) and meta temporal ROI (C&D) and in CU alone (E&F). Clinical groups are colour-coded with red for dementia, green for MCI and blue for CU. Solid circles are Aβ PET positive. The black dashed horizontal line corresponds to the p217+tau threshold derived from Youden’s index. Linear correlation and Spearman co-efficient are shown for the Aβ+ (dark blue) and Aβ-CU (light blue) in part E.

**Figure 5:**
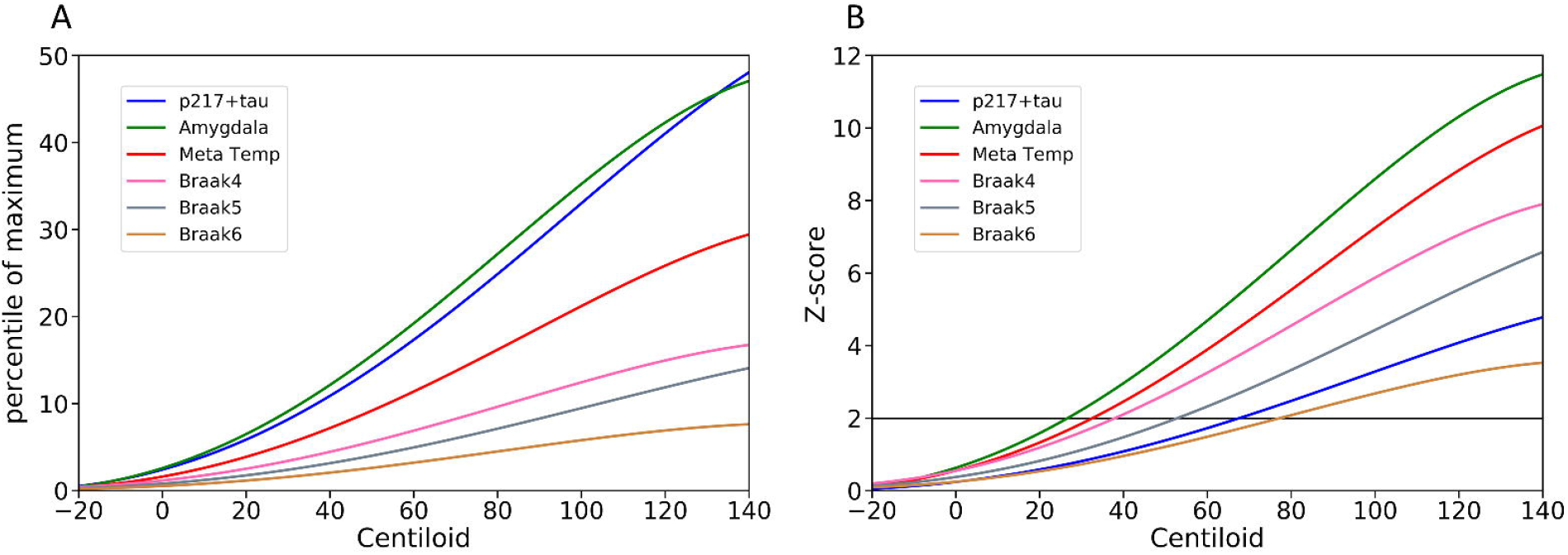
Modelling of ^18^F-MK6240 quantification and p217+tau as a function of CL using polynomial curves; A) normalised by linear transform of p217+tau levels and each tau PET ROI SUVR to a scale of zero to 100 where zero is the mean of the results for each marker in Aβ– CU (<15CL) and 100 is the mean of the results for each marker from the 30 individuals with the highest values for each marker. B) normalised by Z-score using the results from Aβ– CU to define the normal range for p217+tau level and each tau PET ROI SUVR. The horizontal line is +2 S.D. A) suggests that plasma p217+tau rises early, similar to amygdala tau at low levels of Aβ while B) shows that the wide normal range for p217+tau delays reaching a 2 S.D. threshold for significance.

The correlation between p217+tau and tau SUVR was moderate to strong in the meta temporal ROI (Spearman ρ=0.63, p<10^−46^) and the mesial temporal ROI (Me) (Spearman ρ=0.60, p<10^−40^) but was progressively lower when the ROI was restricted to higher Braak stage regions. In CI subjects, the correlation was again strongest in the meta temporal ROI (ρ = 0.69). In CU the correlation was lower; meta temporal ROI (ρ = 0.34); mesial temporal ROI (ρ = 0.33) while Figure 2 shows the regional correlation matches the early and predominant regions of tau accumulation in AD. The correlations persisted between ptau217+ and tau SUVR_MT_ when analysis only included the Aβ+ participants (ρ = 0.52 in CI; ρ = 0.37 in CU). There was no correlation between p217+tau and tau PET in Aβ-individuals (p>0.05). Surface projection of the Pearson correlation available in supplementary Figure 6.

### 4. Performance of plasma p217+tau vs Aβ PET

For discriminating the diagnostic groups, the AUC of p217+tau for Aβ+ AD vs Aβ– CU was 0.94 [0.90-0.97] and for Aβ+ AD vs all CU was 0.88 [0.84-0.92]. For Aβ+ AD vs Aβ– dementia the AUC was 0.93 [0.90-0.96].

For discriminating Aβ+ PET from Aβ– PET in the entire cohort, the AUC was 0.89 [0.86 - 0.93]) or 0.90 if the three Aβ–/tau+ PET individuals (all three had elevated plasma p217+tau) were removed. Youden Index provided a threshold concentration of 126.7fg/ml [100.0– 134.4fg/ml] that yielded an accuracy of 0.85, sensitivity 0.79 [0.75-0.89], specificity 0.89 [0.80-0.92], PPV 0.84, and NPV 0.85. P217+tau accurately identified Aβ+ individuals among CI participants (AUC=0.89, [0.84-0.93]) and among CU (AUC=0.84 [0.76-0.91]). The Youden threshold was also 126.7fg/ml [97.4-177.2] for the CI giving sensitivity 0.82 [0.66-0.92] and specificity 0.82 [0.75-0.98], PPV 0.92 and NPV 0.65. Youden’s index provided an optimal threshold of 100.3fg/ml [100.3-137.3] for CU participants, with a sensitivity of 0.80 [0.78-0.95], specificity of 0.83 [0.67-0.90], PPV 0.49 and NPV 0.95. Applying the whole cohort p217+tau threshold of 126.7fg/ml to CU gave sensitivity of 0.72 [0.6 –0.83], specificity of 0.90 [0.86–0.94], PPV 0.66 and NPV 0.92.

The Z-score threshold for p217+tau derived from CU with <15 CL on Aβ PET was higher at 164 fg/ml resulting in lower sensitivity of 0.60, higher specificity of 0.96, and PPV 0.91 and NPV 0.76 for all subjects to detect positive Aβ PET. In CI, the higher threshold gave sensitivity 0.66, specificity 0.94, PPV 0.96 and NPV 0.53. In CU it gave sensitivity 0.43, specificity 0.96, PPV 0.74 and NPV 0.87.

Altering the CL threshold for Aβ+ PET between 25 to 50 CL had no effect on AUC. However, at 15 and 20 CL thresholds the AUC decreased due to more p217+tau false negative results (Figure 3).

### 5. Performance of plasma p217+tau vs ^18^F-MK6240 tau PET

Plasma p217+tau accurately discriminated individuals with elevated tau in all examined ROIs with the highest AUC in the meta temporal ROI (AUC=0.89 [0.86-0.92]). The scatter plots and ROC curves are shown in Figure 4 for the mesial temporal and meta temporal ROI. The Youden Index threshold was 126.7fg/ml, exactly the same as for Aβ PET and provided an accuracy of 0.81, sensitivity of 0.80 [0.71-0.93] and specificity of 0.82 [0.71-0.93]).

In CU, the highest AUC (0.88 [0.83-0.92]) was observed in the meta temporal region with a Youden’s index threshold of 97.8fm/mL giving sensitivity of 0.9 [0.77-0.97] and specificity of 0.75 [0.71-0.88]. Applying the 126.7fg/ml threshold to the CU group gave sensitivity of 0.71 [0.51-0.84] and specificity of 0.85 [0.80-0.89]. Correlation between p217+tau and tau PET was only significant (p<0.05) in the CU subjects who were Aβ+ and was strongest in the meta temporal ROI (ρ = 0.37) (Figure 4 blue lines).

In CI the highest AUC were observed in the meta temporal and the inferior temporal regions, with AUC’s of 0.86 [0.81-0.91]. The mesial temporal AUC was lower at 0.81 [0.75-0.86]. The Youden’s index threshold for the meta temporal ROI provided a threshold of 148.4fg/mL with a sensitivity of 0.73 [0.65-0.83] and specificity of 0.90 [0.82-0.97]. Applying the threshold of 126.7fg/ml to the CI cohort gave sensitivity of 0.82 [0.77-0.89] and specificity of 0.71 [0.6-0.79].

#### 5.1 p217+tau vs tau PET in subregions of the mesial temporal in cognitively unimpaired

The AUC for p217+tau to detect regional tau+ PET in the CU was higher for the meta temporal region at 0.88 [0.83-0.92] than the amygdala 0.81 [0.74 - 0.88], hippocampus 0.81 [0.74-0.88], parahippocampus 0.86 [0.77-0.93] or entorhinal cortex 0.78 [0.71-0.83]. Plotting of the data points (see supplementary Figure 12) shows that discordance was predominantly due to p217+tau+ve/tau PET-subjects with a disproportionately high prevalence of Aβ+ in this category compared to p217+tau-ve/tau PET-.

### 6. p217+tau predicts Aβ and Tau pathology independent of confounders

AUC values from base models incorporating age, sex and *APOE* ε4 allele status to predict Aβ+ and meta temporal tau+ PET were 0.72 [0.67-0.77] and 0.69 [0.64-0.76] respectively. Adding p217+tau to the base model significantly improved prediction (p-value via DeLong’s ROC test <0.0001) compared to the base model alone and produced AUC of 0.91 [0.86-0.93] for Aβ+ PET and AUC of 0.89 [0.86-0.92] for meta temporal tau+ PET. The combined model, however, did not perform significantly better than p217+tau alone that gave AUC of 0.89 [0.86-0.93] to predict Aβ+ and 0.89 [0.86-0.92] for meta temporal tau+ PET.

### 7. Modelling of ^18^F-MK6240 SUVR and p217+tau as a function of Centiloid

To unveil the chronological order of the emergence of elevation in biomarkers, we normalised ^18^F-MK6240 SUVR and p217+tau concentration to the same scale, using two different methods and mapped against Aβ CL level used as a surrogate for duration of disease development. Firstly, we linearly normalised the range of biomarkers from zero to 100, with zero being the mean of the CU Aβ-individuals with less than 15 CL to increase certainty of Aβ negativity [24], and 100 being the mean of the 30 individuals with the highest biomarker value. When normalising the range of values between 0 and 100, (Figure 5A), the fitted polynomial curves of ^18^F-MK6240 in amygdala and plasma p217+tau were nearly identical and rose very early when Aβ level was quite low and before other tau PET regions began to rise. Secondly, we normalized each biomarker to a Z-score, again using those CU with <15 CL to provide the normal range. Figure 5B shows that although we have demonstrated in Figure 5A that plasma p217+tau rises early, it takes until 70CL to exceed two standard deviations of the normal range.

## Discussion

We determined the performance of a plasma ptau assay that utilizes an antibody raised against the paired helical filaments (PHF) of Alzheimer’s disease. Human PHF tau is phosphorylated at multiple sites [27]. Plasma p217+tau measures tau phosphorylated at aa217 with binding enhanced by the simultaneous presence of phosphorylation at aa212 so may better reflect the tau of AD than single binding site ptau assays [12]. To evaluate this assay, we employed PET tracers that may give greater sensitivity than those previously used for comparison to plasma ptau [7, 21, 28].

Plasma p217+tau displayed high accuracy in detecting Aβ+ individuals across the clinical spectrum. Compared to Aβ– PET CU, plasma p217+tau concentration was two-fold higher in Aβ+ CU and 3.5-fold higher in Aβ+ MCI and dementia. The AUC for Aβ+AD vs Aβ-CU was 0.94, for Aβ+AD vs Aβ– dementia was 0.93 while the AUC for all Aβ+ vs Aβ- and for tau+ vs tau-were equal at 0.89. In the CU group, the AUC was 0.84 for Aβ+ vs Aβ-PET and 0.88 for tau+ vs tau-PET. Adding age, sex and *APOE*ε4 did not significantly improve the prediction of Aβ+ PET (AUC 0.89 vs 0.91) or tau+ PET (unchanged at 0.89). These results compare favourably to other plasma ptau assays though the use of different assay methods, different anti-tau antibodies, and cohort variation prevent direct comparison. The range of AUC reported for Aβ+ vs Aβ-PET for plasma ptau measures has ranged from 0.76 to 0.92 [2, 3, 7-9, 29, 30]. Our results with p217+tau are at the high end of reported plasma ptau assay performance and used Simoa technology that is kit based, fully automated and has a large installation base and therefore is well suited to widespread deployment.

Our data (Figures 3-5) suggest plasma p217+tau begins to rise i) soon after brain Aβ levels begin to trend up as assessed by PET, ii) concordant with the rise in tau in the amygdala region on PET, and iii) prior to the rise in meta temporal tau PET. This is supported by the vertex-based analysis of correlation in CU, that p217+tau and Aβ are highly associated in brain regions where early Aβ deposition occurs and with tau in the anteromedial temporal lobe (Figure 2). However, the plasma p217+tau level does not reach +2 standard deviations above the mean found in Aβ-CU until Aβ reaches moderate levels (approximately 70 CL). These findings are consistent with reports for plasma ptau measures of p181, p217 and p231 [6, 9, 31, 32]. Wide intersubject variability in plasma ptau in persons without evidence of AD pathology on amyloid PET remains a challenge and warrants further investigation.

Our data also suggests that tau aggregates in the amygdala region and plasma p217+tau increase together and very early in the development of AD when Aβ load is beginning to rise (Figure 5A). The amygdala performed best in our comparison of p217+tau with tau PET mesial temporal subregions. This may be due to reduced partial volume effect compared to smaller structures and less spill-over effect from off-target binding in the meninges giving a more robust signal. The effect of primary age related tauopathy (PART) where tau aggregates are present without Aβ plaques [33] and predominantly in the entorhinal cortex, on plasma p217+tau levels is unclear and requires further investigation.

Only longitudinal studies will determine if what appear to be false positive p217+tau results in CU compared to PET are due to variation and limitation in the plasma measure or are detecting very early stage AD prior to PET becoming significantly positive and this work is in progress under the AIBL study umbrella. In contrast to the CU, the CI show more “false negative” p217+tau results when compared to both Aβ and tau PET. Many of these false negative p217+tau results were A+/T+ on PET so this requires further investigation.

Reflecting the impact of disease prevalence in our study population (71% Aβ+ in CI vs 21% in CU), the p217+tau positive and negative predictive values of p217+tau for Aβ+ PET were very different for the CI and CU cohorts. The PPV were high in the CI group for predicting Aβ (0.94) and tau (0.93), suggesting that p217+tau may be a good test to confirm AD in patients with objective cognitive impairment. However, the NPV were relatively low at 0.66 and 0.65 suggesting that a negative p217+tau does not exclude AD. Lowering the p217+tau threshold to 100 fg/ml in the CI group only improved the NPV to 0.73 while lowering PPV to 0.88 and at 75 fg/ml the NPV was 0.78, PPV 0.84. P217+tau has potential as a diagnostic tool and combination with other biomarkers, as discussed by Schindler et al [34], may increase accuracy and provide an affordable and accurate diagnostic tool. Conversely, in the CU group the NPV were high (>0.94), suggesting that a negative p217+tau test can largely exclude AD pathology in persons with normal cognitive test results. However, the associated lower PPV (0.50) in CU indicates that positive results would need confirmation by PET or CSF analysis. These findings show that triage with p217+tau may substantially reduce the number of screening Aβ PET needed to identify Aβ+ PET participants for preclinical AD trials resulting in considerable recruitment cost savings.

This study has several limitations. First the results need validation in other cohorts and more diverse populations. Secondly this is a cross-sectional study and longitudinal analysis is required to confirm the predictive value of p217+tau as a marker of disease progression and to clearly determine when abnormal levels of p217+tau can be measured along the disease trajectory.

In conclusion, an elevated level of plasma p217+tau is associated with both elevated Aβ and tau across the clinical spectrum of Alzheimer’s disease. Elevated p217+tau strongly supports a diagnosis of AD in persons with MCI or dementia while a low level in cognitively unimpaired persons is strong evidence against preclinical AD.

## Data Availability

The datasets used and/or analysed during the current study are available from the corresponding author on reasonable request.

## Declarations

### Ethics approval and Consent to participate

This study was approved by the Austin Health Human Research Ethics Committee (HREC/18/Austin/201)

### Consent for publication

All participants gave written consent for publication of de-identified data.

## Acknowledgement

Some of the data used in the preparation of this article was obtained from the Australian Imaging Biomarkers and Lifestyle flagship study of aging (AIBL), funded by the Commonwealth Scientific and Industrial Research Organization (CSIRO), National Health and Medical Research Council (NHMRC), and participating institutions. AIBL researchers are listed at www.aibl.csiro.au. The authors thank all participants who took part in the study, as well as their families.

