## Supplementary material for "Plasma p217+tau vs NAV4694 amyloid and MK6240 tau PET across the Alzheimer continuum"

|  | **CU Aβ-**  (n=177) | **CU Aβ+**  (n=46) | **MCI Aβ-**  (n=35) | **MCI Aβ+**  (n=56) | **Dem. Aβ-**  (n=15) | **AD Aβ+**  (n=68) |
| --- | --- | --- | --- | --- | --- | --- |
| **Age** | 74.6 (5.3) | 77.6 (6.5) | 70.0 (8.6) | 75.9 (6.7) | 71.1 (6.5) | 70.6 (8.2) |
| **Gender N male (% )** | 83 (47%) | 18 (39%) | 19 (54%) | 32 (57%) | 10 (67%) | 39 (57%) |
| **Education (years)** | 14.2 (3.0) | 12.9 (2.8) | 12.5 (3.5) | 12.4 (2.9) | 10.8 (3.6) | 12.2 (2.8) |
| **APOE ε4 (%)** | 45 (25%) | 27 (59%) | 7 (20%) | 39 (70%) | 2 (13%) | 44 (65%) |
| **MMSE median (IQR)** | 29.0 (2) | 28.0 (2) | 28.0 (2) | 26.0 (3) | 24.0 (4.5) | 23.0 (4) |
| **CDR SoB median (IQR)** | 0.0 (0.0) | 0.0 (0.0) | 0.5 (0.5) | 1.5 (1.0) | 4.5 (1.2) | 4.0 (1.1) |
| **AIBL PACCs mean (SD)** | -0.33 (0.76) | -0.60 (1.10) | -1.74 (1.08) | -2.77 (0.96) | -3.62 (1.26) | -4.48 (2.55) |
| **Centiloid mean (SD)** | 3.1 (7.8) | 84.5 (39.4) | 1.5 (5.8) | 117.3 (43.5) | 4.8 (7.5) | 112.4 (40.5) |
| **tau SUVR_MT_ mean (SD)** | 0.96 (0.12) | 1.25 (0.35) | 1.07 (0.45) | 1.74 (0.56) | 1.09 (0.27) | 2.11 (0.73) |
| **tau SUVR_MT_+ N (%)** | 9 (5%) | 22 (48%) | 2 (6%) | 46 (82%) | 3 (20%) | 61 (90%) |
| **Adjusted HV** | 2.99 (0.26) | 2.93 (0.25) | 2.92 (0.41) | 2.66 (0.35) | 2.54 (0.60) | 2.58 (0.38) |
| **Plasma p217+tau** | 71.2 (46.6) | 162.7 (85.8) | 82.1 (55.8) | 247.1 (141.2) | 93.4 (45.6) | 260.5 (147.7) |

Supplementary Table 1: Cohort demographics split by diagnostic and Aβ status

**Analysis using 126.7fg/mL threshold**

| **Centiloid** | **AUC** | **Accuracy** | **Sensitivity** | **Specificity** | **PPV** | **NPV** | **Threshold** |
| --- | --- | --- | --- | --- | --- | --- | --- |
| **15CL** | 0.78  (0.71-0.86) | 0.8 | 0.55 | 0.91 | 0.7 | 0.83 | 126.7 |
| **20CL** | 0.8  (0.72-0.87) | 0.83 | 0.6 | 0.9 | 0.68 | 0.87 | 126.7 |
| **25CL** | 0.85  (0.78-0.92) | 0.87 | 0.72 | 0.9 | 0.66 | 0.92 | 126.7 |
| **30CL** | 0.85  (0.78-0.93) | 0.86 | 0.72 | 0.89 | 0.62 | 0.93 | 126.7 |
| **40CL** | 0.87  (0.79-0.94) | 0.86 | 0.74 | 0.88 | 0.56 | 0.94 | 126.7 |
| **50CL** | 0.89  (0.82-0.95) | 0.85 | 0.76 | 0.87 | 0.5 | 0.95 | 126.7 |

Supplementary Table 2 Statistics of the ROC analysis to predict amyloid positivity in CU for different CL threshold using a p217+tau threshold of 126.7fg/mL.

| **Centiloid** | **AUC** | **Accuracy** | **Sensitivity** | **Specificity** | **PPV** | **NPV** | **Threshold** |
| --- | --- | --- | --- | --- | --- | --- | --- |
| **15CL** | 0.89  (0.84–0.94) | 0.82 | 0.82 | 0.84 | 0.93 | 0.64 | 126.7 |
| **20CL** | 0.89  (0.83-0.94) | 0.82 | 0.81 | 0.82 | 0.92 | 0.64 | 126.7 |
| **25CL** | 0.89  (0.83-0.94) | 0.82 | 0.81 | 0.82 | 0.92 | 0.64 | 126.7 |
| **30CL** | **0.90**  **(0.85-0.95)** | 0.83 | 0.83 | 0.83 | 0.92 | 0.67 | 126.7 |
| **40CL** | 0.89  (0.84-0.94) | 0.82 | 0.83 | 0.79 | 0.89 | 0.69 | 126.7 |
| **50CL** | 0.88  (0.83-0.93) | 0.8 | 0.83 | 0.74 | 0.85 | 0.7 | 126.7 |

Supplementary Table 3 Statistics of the ROC analysis to predict amyloid positivity in CI for different CL threshold using a p217+tau threshold of 126.7fg/mL.

**Analysis using the Youden’s index**

| **Centiloid** | AUC | Accuracy | Sensitivity | Specificity | PPV | NPV | Threshold |
| --- | --- | --- | --- | --- | --- | --- | --- |
| **15CL** | 0.87  (0.83-0.90) | 0.81 | 0.81 | 0.81 | 0.8 | 0.83 | 104.5 |
| **20CL** | 0.87  (0.84-0.90) | 0.82 | 0.75 | 0.88 | 0.84 | 0.81 | 126.7 |
| **25CL** | 0.89  (0.86-0.93) | 0.85 | 0.79 | 0.89 | 0.84 | 0.85 | 126.7 |
| **30CL** | 0.9  (0.87-0.93) | **0.85** | 0.81 | 0.88 | 0.82 | 0.86 | 126.7 |
| **40CL** | **0.90**  (0.87-0.93) | 0.85 | 0.81 | 0.86 | 0.79 | 0.88 | 126.7 |
| **50CL** | 0.90  (0.86-0.92) | 0.84 | 0.82 | 0.84 | 0.74 | 0.89 | 126.7 |

Supplementary Table 4 Statistics of the ROC analysis to predict amyloid positivity the whole cohort for different CL threshold using Youden’s index.

| **Centiloid** | **AUC** | **Accuracy** | **Sensitivity** | **Specificity** | **PPV** | **NPV** | **Threshold** |
| --- | --- | --- | --- | --- | --- | --- | --- |
| **15CL** | 0.78  (0.71-0.86) | 0.79 | 0.7 | 0.82 | 0.61 | 0.87 | 100.33 |
| **20CL** | 0.8  (0.72-0.87) | 0.79 | 0.74 | 0.81 | 0.57 | 0.9 | 100.33 |
| **25CL** | 0.85  (0.78-0.92) | 0.81 | 0.83 | 0.8 | 0.51 | 0.95 | 100.33 |
| **30CL** | 0.85  (0.78-0.93) | 0.8 | 0.84 | 0.79 | 0.49 | 0.95 | 100.33 |
| **40CL** | 0.87  (0.79-0.94) | 0.8 | 0.87 | 0.78 | 0.44 | 0.97 | 100.33 |
| **50CL** | 0.89  (0.82-0.95) | 0.8 | 0.88 | 0.79 | 0.41 | 0.97 | 105.16 |

Supplementary Table 5 Statistics of the ROC analysis to predict amyloid positivity in CU for different CL threshold using Youden’s index.

| **Centiloid** | **AUC** | **Accuracy** | **Sensitivity** | **Specificity** | **PPV** | **NPV** | **Threshold** |
| --- | --- | --- | --- | --- | --- | --- | --- |
| **15CL** | 0.89  (0.84–0.94) | 0.83 | 0.82 | 0.84 | 0.93 | 0.65 | 126.68 |
| **20CL** | 0.89  (0.83-0.94) | 0.82 | 0.82 | 0.82 | 0.92 | 0.65 | 126.68 |
| **25CL** | 0.89  (0.83-0.94) | 0.82 | 0.82 | 0.82 | 0.92 | 0.65 | 126.68 |
| **30CL** | **0.90**  **(0.85-0.95)** | **0.83** | **0.84** | **0.83** | **0.92** | **0.68** | **126.68** |
| **40CL** | 0.89  (0.84-0.94) | 0.82 | 0.84 | 0.79 | 0.89 | 0.7 | 126.68 |
| **50CL** | 0.88  (0.83-0.93) | 0.79 | 0.73 | 0.9 | 0.93 | 0.64 | 148.41 |

Supplementary Table 6 Statistics of the ROC analysis to predict amyloid positivity in MCI and AD for different CL threshold using Youden’s index.

| **Region** | **AUC**  **(95%CI)** | **Accuracy** | **Sensitivity** | **Specificity** | **PPV** | **NPV** | **Threshold**  **(fg/ml)** |
| --- | --- | --- | --- | --- | --- | --- | --- |
| **Mesial Temporal** | 0.85  (0.81-0.88) | 0.78 | 0.75 | 0.79 | 0.67 | 0.85 | 126.7 |
| **Meta Temporal** | 0.89  (0.86-0.92) | 0.81 | 0.8 | 0.82 | 0.71 | 0.88 | 126.7 |
| **Inf. Temporal** | 0.86  (0.83-0.90) | 0.85 | 0.70 | 0.92 | 0.80 | 0.87 | 173.2 |
| **Braak IV** | 0.87  (0.83-0.90) | 0.86 | 0.72 | 0.91 | 0.76 | 0.89 | 173.2 |
| **Braak V** | 0.87  (0.83-0.90) | 0.86 | 0.7 | 0.91 | 0.74 | 0.90 | 186.0 |
| **Braak VI** | 0.82  (0.77-0.86) | 0.83 | 0.69 | 0.86 | 0.54 | 0.92 | 186.0 |

Supplementary Table 7: Results from the ROC analyses to predict tau positivity in separate brain regions with Youden’s index. Positive predictive value (PPV), Negative predictive value (NPV).

| **Region** | **AUC**  **(95%CI)** | **Accuracy** | **Sensitivity** | **Specificity** | **PPV** | **NPV** | **Threshold**  **(fg/ml)** |
| --- | --- | --- | --- | --- | --- | --- | --- |
| **Mesial Temporal** | 0.8  (0.72-0.87) | 0.74 | 0.81 | 0.73 | 0.32 | 0.96 | 97.8 |
| **Meta Temporal** | 0.88  (0.83-0.92) | 0.77 | 0.9 | 0.75 | 0.36 | 0.98 | 97.8 |
| **Inf. Temporal** | 0.75  (0.65-0.84) | 0.7 | 0.73 | 0.7 | 0.2 | 0.96 | 97.8 |
| **Braak IV** | 0.82  (0.72-0.90) | 0.73 | 0.81 | 0.72 | 0.17 | 0.98 | 104.5 |
| **Braak V** | 0.75  (0.61-0.87) | 0.72 | 0.75 | 0.72 | 0.16 | 0.97 | 104.5 |
| **Braak VI** | 0.66  (0.49-0.81) | 0.86 | 0.45 | 0.88 | 0.13 | 0.97 | 151.8 |

Supplementary Table 8: Results from the ROC analyses to predict tau positivity in CU and in different brain regions with Youden’s index. Positive predictive value (PPV), Negative predictive value (NPV).

| **Region** | **AUC**  **(95%CI)** | **Accuracy** | **Sensitivity** | **Specificity** | **PPV** | **NPV** | **Threshold**  **(fg/ml)** |
| --- | --- | --- | --- | --- | --- | --- | --- |
| **Mesial Temporal** | 0.81  (0.75–0.86) | 0.73 | 0.65 | 0.90 | 0.92 | 0.57 | 190.8 |
| **Meta Temporal** | 0.86  (0.81-0.91) | 0.79 | 0.73 | 0.90 | 0.93 | 0.65 | 148.4 |
| **Inf. Temporal** | 0.86  (0.81-0.9) | 0.82 | 0.77 | 0.89 | 0.91 | 0.74 | 161.0 |
| **Braak IV** | 0.83  (0.78-0.88) | 0.79 | 0.74 | 0.84 | 0.86 | 0.72 | 173.2 |
| **Braak V** | 0.83  (0.78-0.88) | 0.78 | 0.74 | 0.84 | 0.82 | 0.75 | 190.8 |
| **Braak VI** | 0.77  (0.71-0.83) | 0.73 | 0.70 | 0.76 | 0.63 | 0.80 | 212.7 |

Supplementary Table 9: Results from the ROC analyses to predict tau positivity in CI and in different brain regions with Youden’s index. Positive predictive value (PPV), Negative predictive value (NPV).

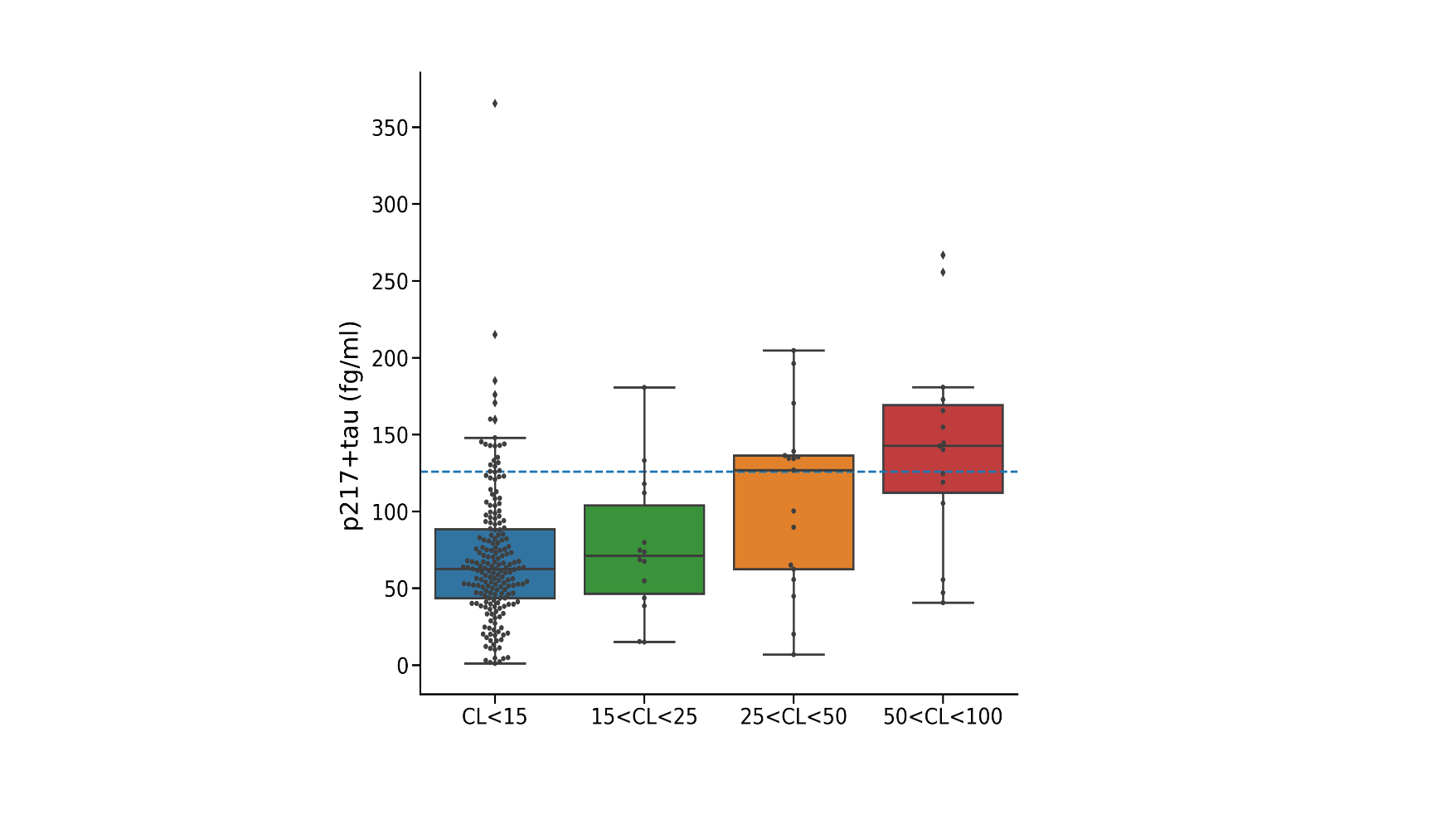

Supplementary Figure 1 Plasma p217+tau concentrations for different level of Aβ in individuals with a negative tau scan. The dash blue line corresponds to the threshold from the Youden’s index that best discriminate Aβ- from Aβ+ individuals.

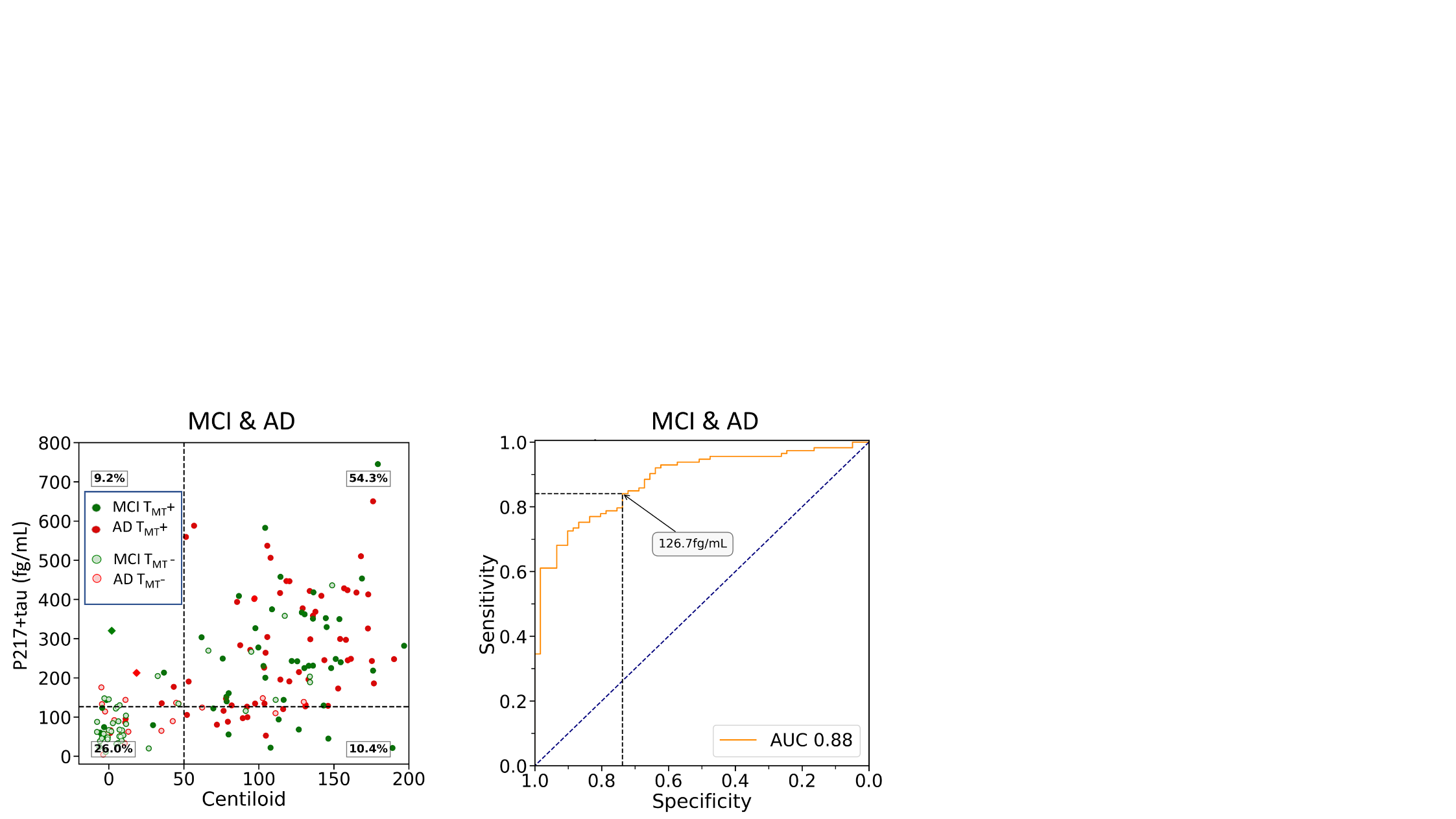

Supplementary Figure 2 On the left, scatter plots of Centiloid versus the plasma p217+tau in 2 different color-coded clinical diagnostic groups (MCI and AD), with light colours used to identify T- subjects. Thresholds are displayed in fine dash vertical and horizontal lines. The dash black vertical line corresponds to the Youden’s index and the blue one to the 85% of specificity at 50CL. The diamond shapes identified the 3 subjects with a suspicious Aβ- PET scan. The % of individuals in the 4 groups A-tau- A+ptau- A-ptau+ A+ptau+ are superimposed to the image. The figure on the right displays the ROC curve.

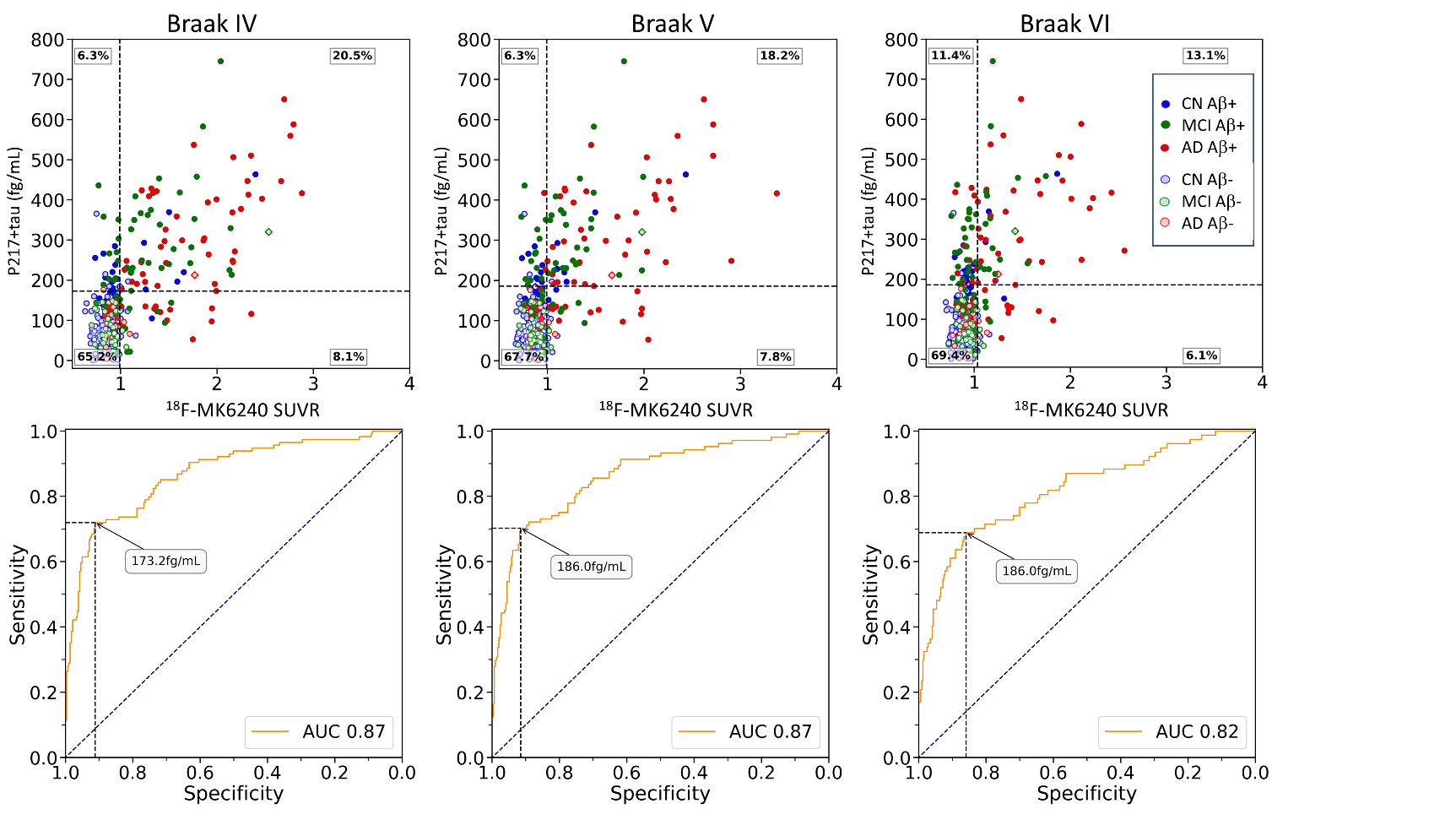

Supplementary Figure 3 Scatter plot of the plasma concentration of p217+tau versus the composite MK6240 SUVR versus the P-tau217 in 3 different color-coded clinical diagnostic groups, with light colours used to identify A- subjects (CL<25). Thresholds are displayed in fine dash vertical and horizontal lines. The dash black vertical line corresponds to the Youden’s index and the blue one to the 85% of specificity.

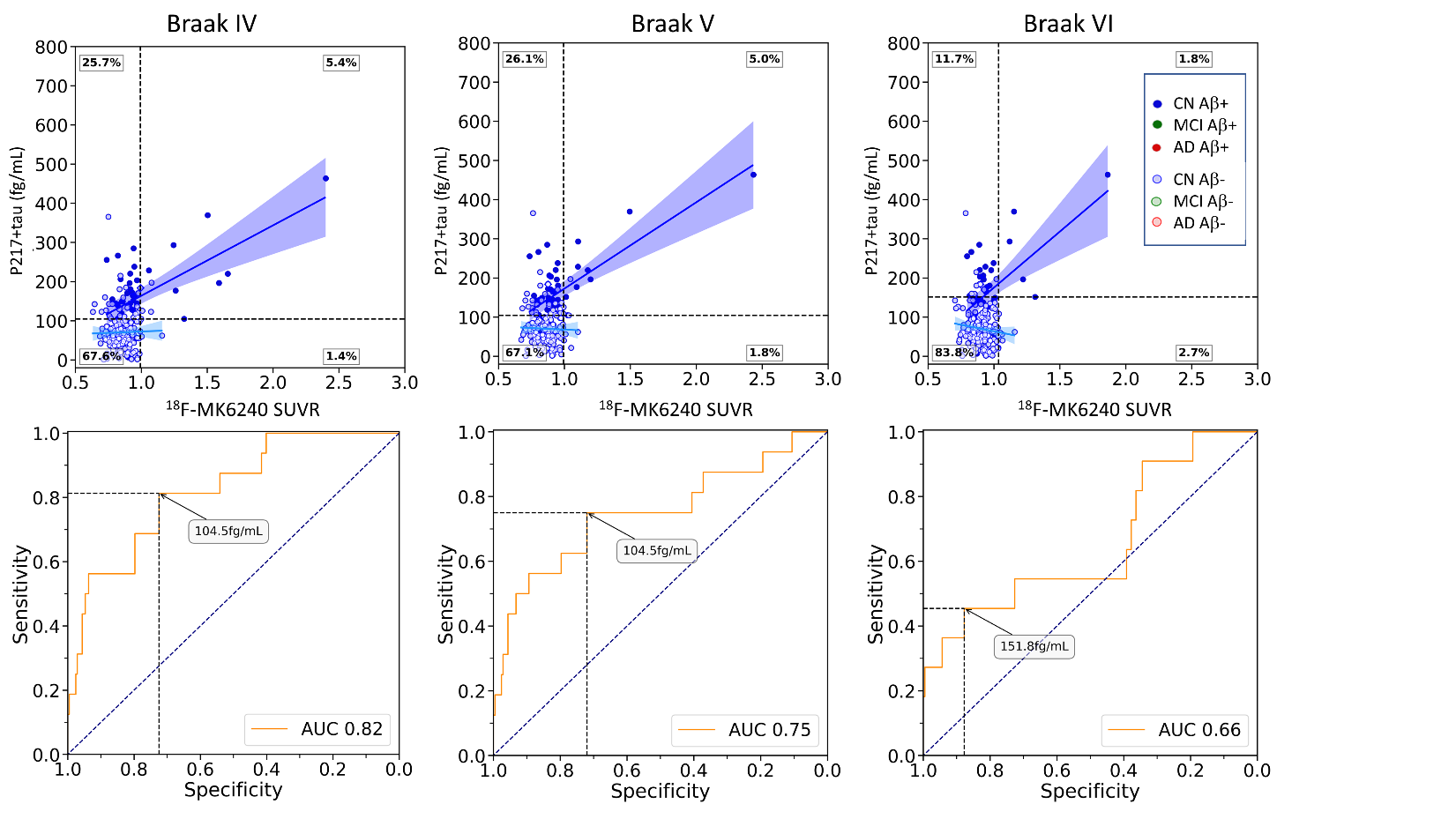

Supplementary Figure 4 Scatter plots of plasma p217+tau concentration versus composite MK6240 SUVR in cognitively unimpaired individuals, with light colours used to identify Aβ- subjects (CL<25). Thresholds are displayed in fine dash vertical and horizontal lines. The dash black vertical line corresponds to the Youden’s index and the blue one to the 85% of sensitivity. The diamond shape identified the individual with a suspicious Aβ- PET scan. The % of individuals in the 4 groups T-p217- T+p217- T-p217+ T+p217+ (Youden) are superimposed to the image. Bottom row, ROC curve, p217+tau predicting regional PET tau status in CU.

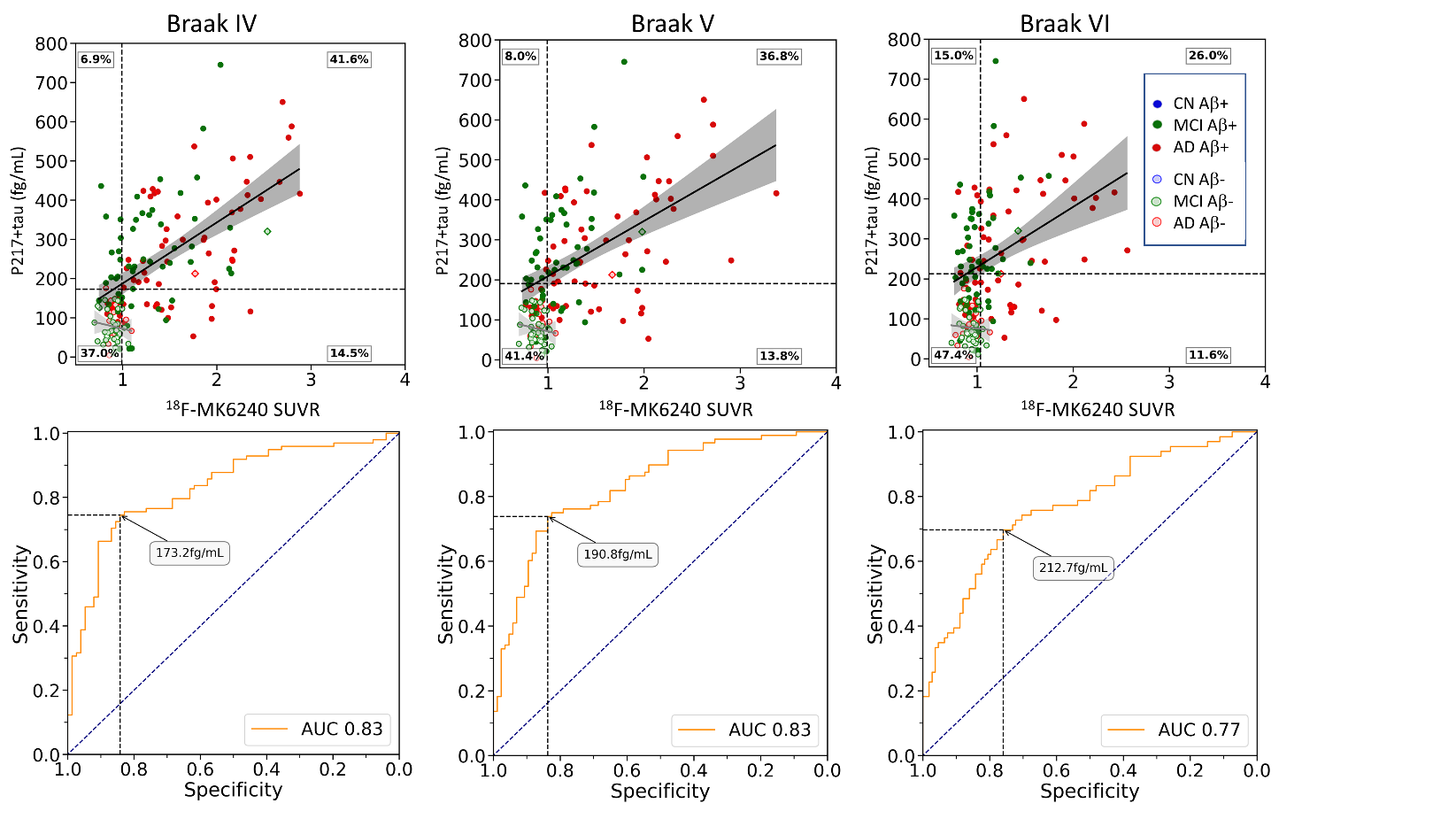

Supplementary Figure 5 Scatter plots of plasma p217+tau concentration versus composite MK6240 SUVR in cognitively unimpaired individuals, with light colours used to identify A- subjects (CL<25). Thresholds are displayed in fine dash vertical and horizontal lines. The dash black vertical line corresponds to the Youden’s index and the blue one to the 85% of sensitivity. The diamond shape identified the 2 individuals with a suspicious Aβ- PET scan. The % of individuals in the 4 groups T-p217- T+p217- T-p217+ T+p217+ (Youden) are superimposed to the image. Bottom row, ROC curve, p217+tau predicting regional PET tau status in CI.

|  | **All** | **CU** | **CI** |
| --- | --- | --- | --- |
| Mesial Temporal | 0.60* | 0.33* | 0.60* |
| Meta Temporal | 0.63* | 0.34* | 0.69* |
| Inf. Temp | 0.55* | 0.21* | 0.66* |
| Braak IV | 0.54* | 0.22* | 0.63* |
| Braak V | 0.47* | 0.14 | 0.57* |
| Braak VI | 0.31* | 0.02 | 0.41* |

Supplementary Table 10 The Spearman rank-order correlation between MK6240 SUVR estimated in several composite ROIs and the concentration of plasma p217+tau. *(p<0.05, corrected for multiple comparison)

|  | **CU Aβ−** | **CU Aβ+** | **CI Aβ−** | **CI Aβ+** |
| --- | --- | --- | --- | --- |
| Mesial Temporal | 0.12 | 0.24 | 0.19 | 0.38* |
| Meta Temporal | 0.11 | 0.37* | 0.16 | 0.52* |
| Inf. Temporal | 0.03 | 0.39* | 0.08 | 0.54* |
| Braak IV | 0.07 | 0.37* | 0.08 | 0.51* |
| Braak V | -0.01 | 0.36* | 0.06 | 0.48* |
| Braak VI | -0.08 | 0.25 | 0.08 | 0.36* |

Supplementary Table 11 The Spearman rank-order correlation between MK6240 SUVR estimated in several composite ROIs and the concentration of plasma p217+tau, *(p<0..05, corrected for multiple comparison)

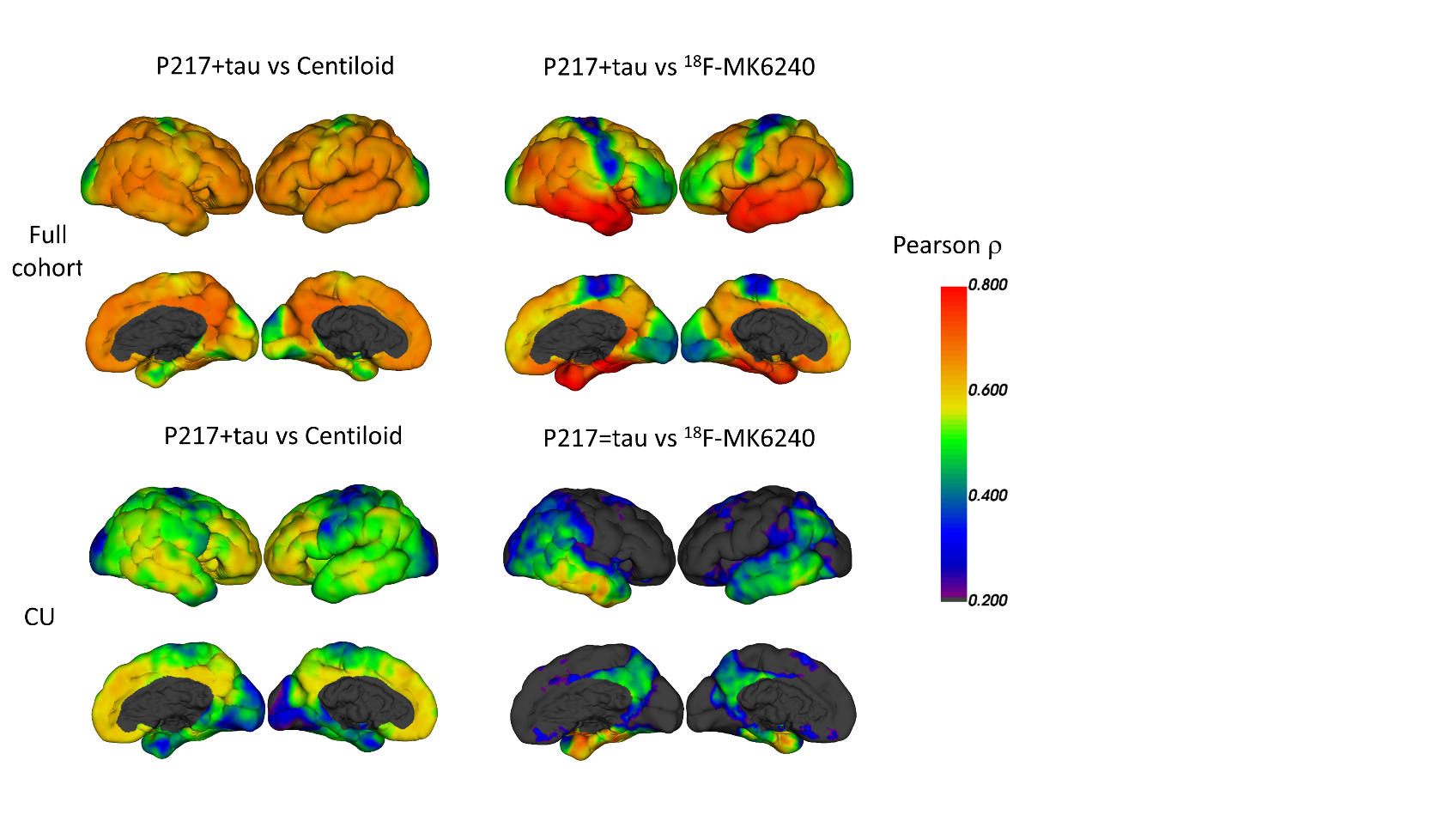

Supplementary Figure 6 Pearson correlations between Ptau-217 and Centiloid (raw1&2) and MK6240 (raw 3 &4). The correlation was for the whole population row1&3 and for HC (raw 2 &4)

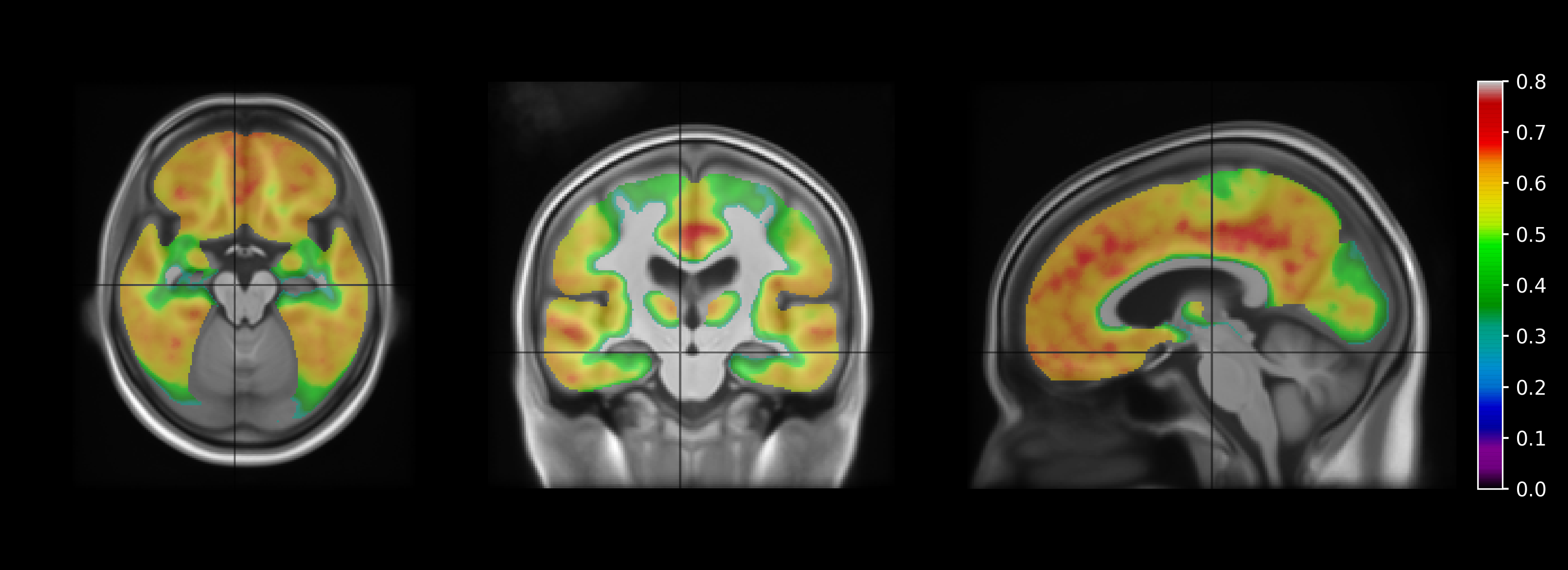

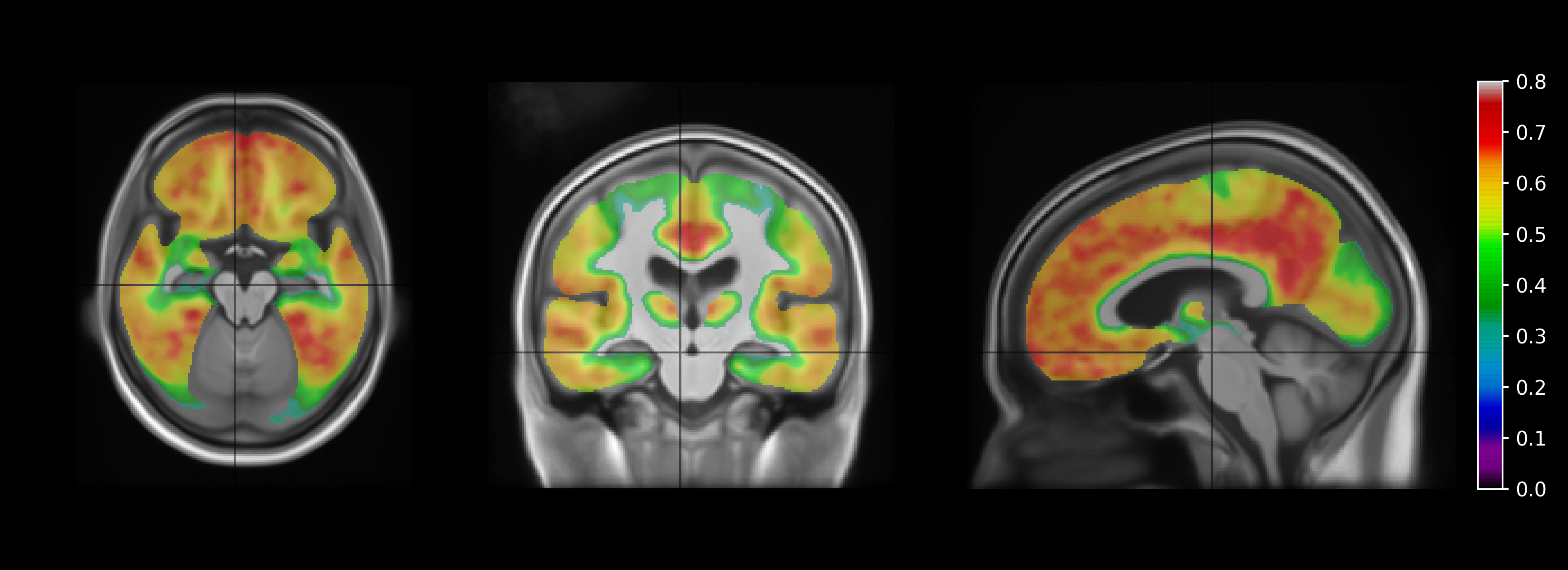

Supplementary Figure 7 VBM analysis of the association between Centiloid and p217+tau (Spearman ρ on top and Pearson **ρ** on bottom)

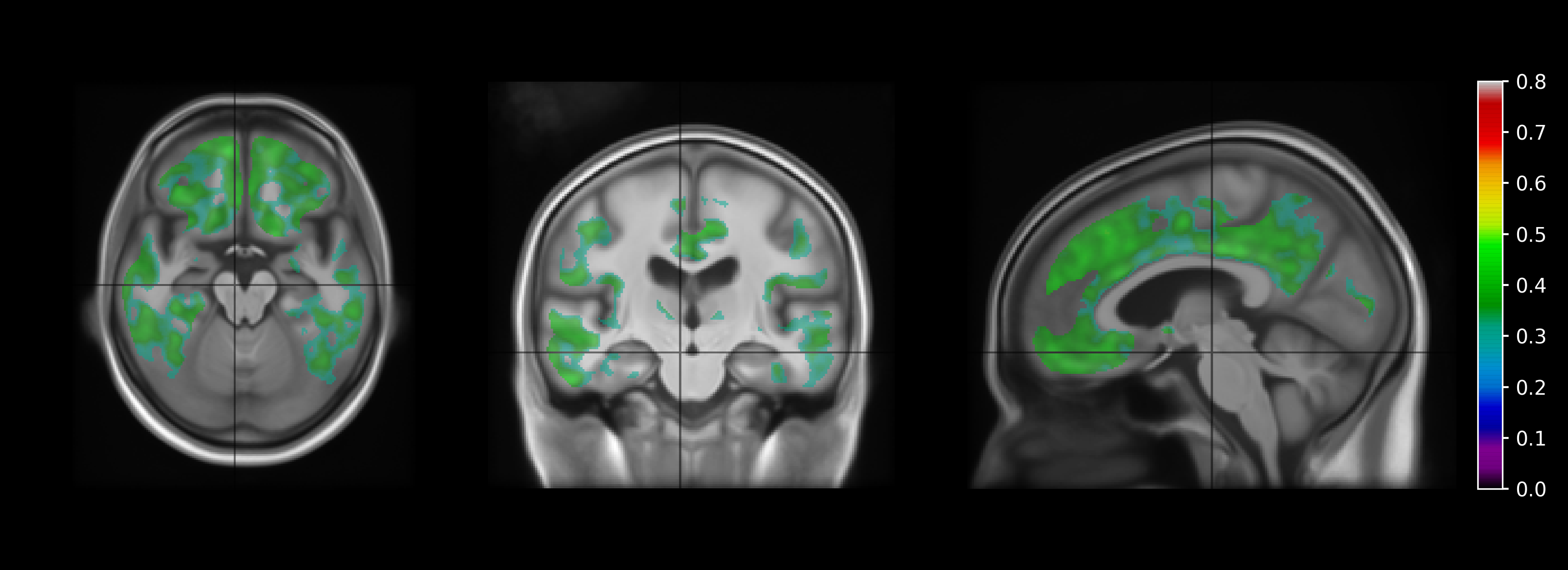

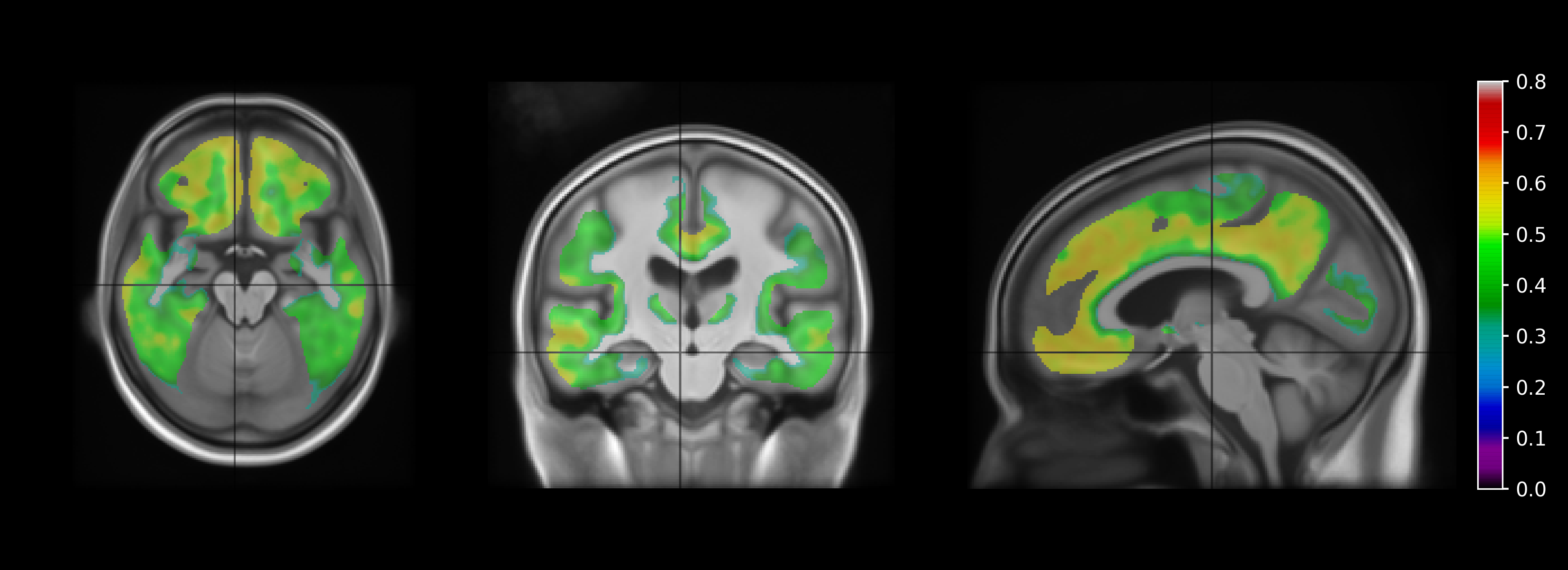

Supplementary Figure 8 VBM analysis of the association between Centiloid and p217+tau in CU (Spearman ρ on top and Pearson **ρ** on bottom)

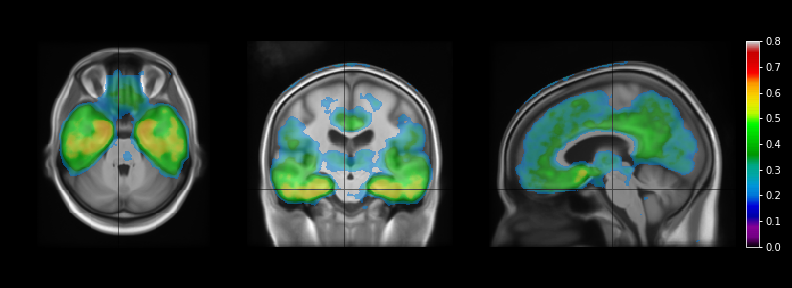

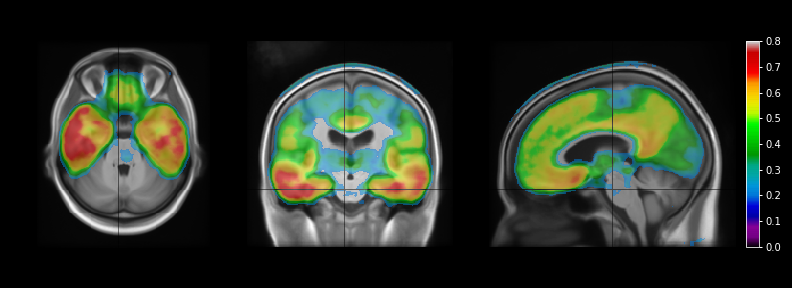

Supplementary Figure 9 VBM analysis of the association between ^18^F-MK6240 SUVR and p217+tau (Spearman ρ on top and Pearson **ρ** on bottom)

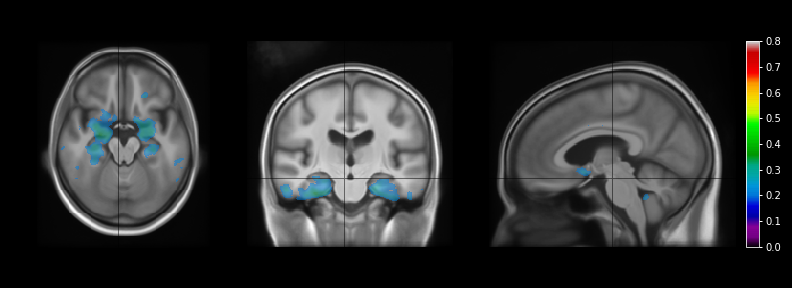

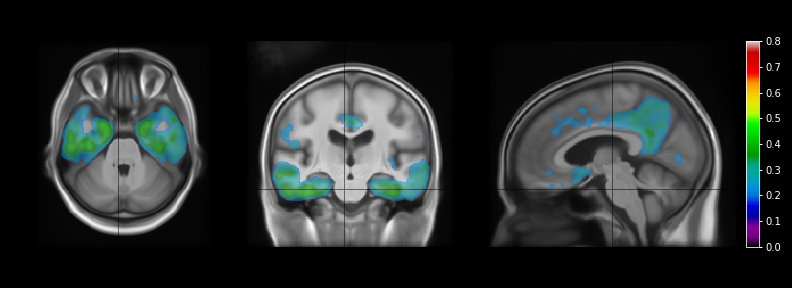

Supplementary Figure 10 VBM analysis of the association between 18F-MK6240 SUVR and p217+tau (Spearman ρ on top and Pearson **ρ** on bottom)

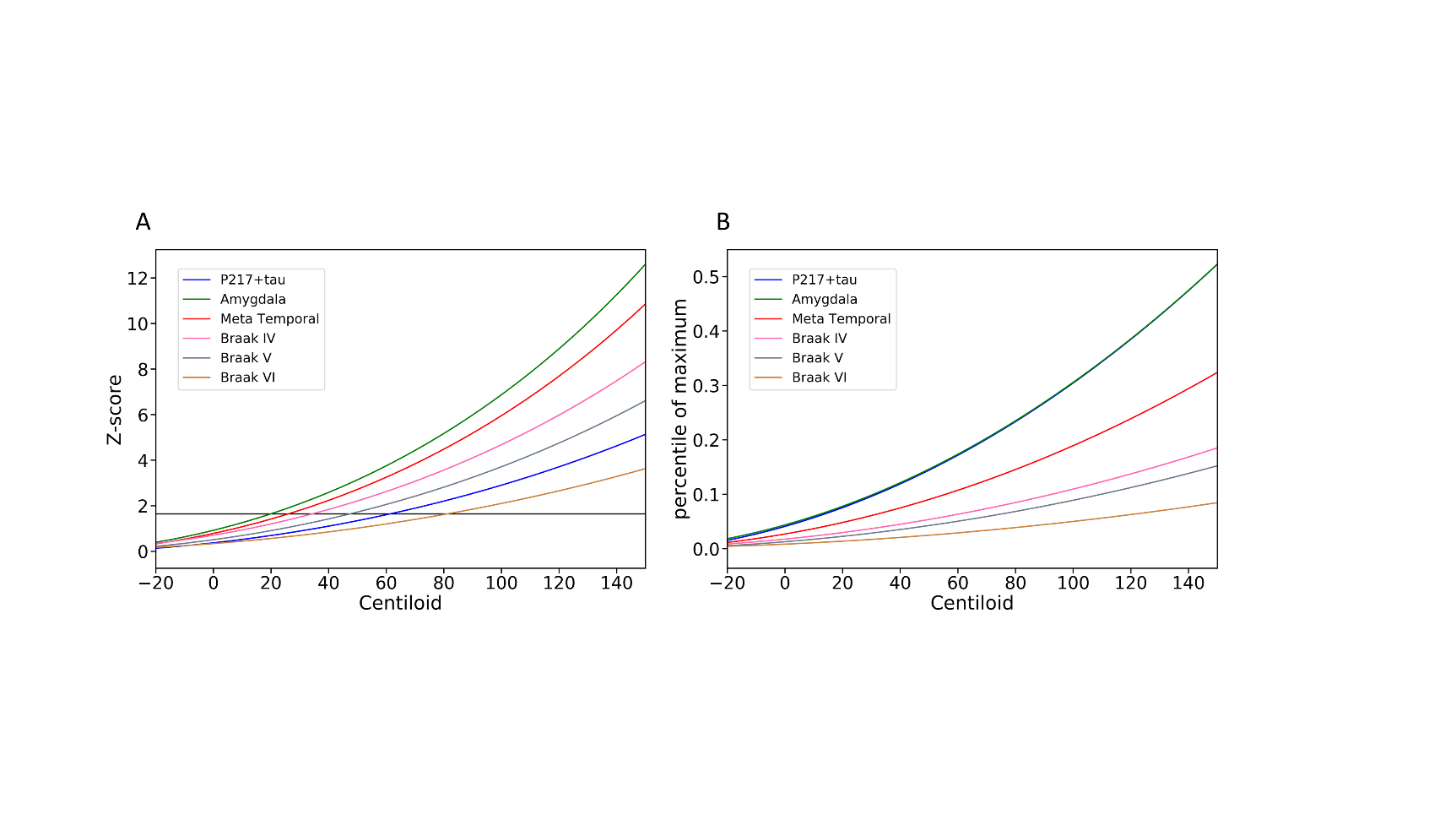

Supplementary Figure 11 Modelisation of ^18^F-MK6240 quantification and p217+tau as a function of Centiloid using exponential fitting, Z-score normalisation on the right (using CU Aβ<15CL to define normal range) and maximum range normalisation (using the top 30 highest p217+tau and SUVR to scale the curves)

**Accuracy of p217+tau in detecting tau in early regions of tau aggregation**

In the cognitive unimpaired group, the predictive values of p217+tau was high in the amygdala (AUC=0.81, [95%CI: 0.74 - 0.88]), parahippocampus (AUC=0.86 [95%CI: 0.77 - 0.93]) and hippocampus (AUC=0.81, [95%CI: 0.74 - 0.88]) and lower in the entorhinal cortex (AUC=0.78 [95%CI: 0.71 - 0.83]). However, there were less participants with high tau uptake in the parahippocampus (6.7%) and in the hippocampus (11.8%) than in the entorhinal cortex (23.9%) and the amygdala (14.4%), where tau may start accumulating earlier. The AUC is these sub-regions were of the same order than the global mesial Me ROI but lower than the Meta temporal region.

| **Region** | **AUC**  **(95%CI)** | **Accuracy** | **Sensitivity** | **Specificity** | **PPV** | **NPV** | **Threshold** |
| --- | --- | --- | --- | --- | --- | --- | --- |
| Entorhinal cortex | 0.78  (0.71-0.83) | 0.66 | 0.87 | 0.59 | 0.40 | 0.93 | 69.5 |
| Amygdala | 0.81  (0.74-0.88) | 0.77 | 0.76 | 0.77 | 0.35 | 0.95 | 105.2 |
| Parahippocampus | 0.86  (0.77-0.93) | 0.8 | 0.8 | 0.8 | 0.21 | 0.98 | 123.2 |
| Hippocampus | 0.81  (0.74-0.88) | 0.73 | 0.81 | 0.72 | 0.28 | 0.97 | 97.8 |

Supplementary Table 12: Results from the ROC analysis to predict tau positivity in CI and in early region of tau deposition with Youden’s index. Positive predictive value (PPV), Negative predictive value (NPV)

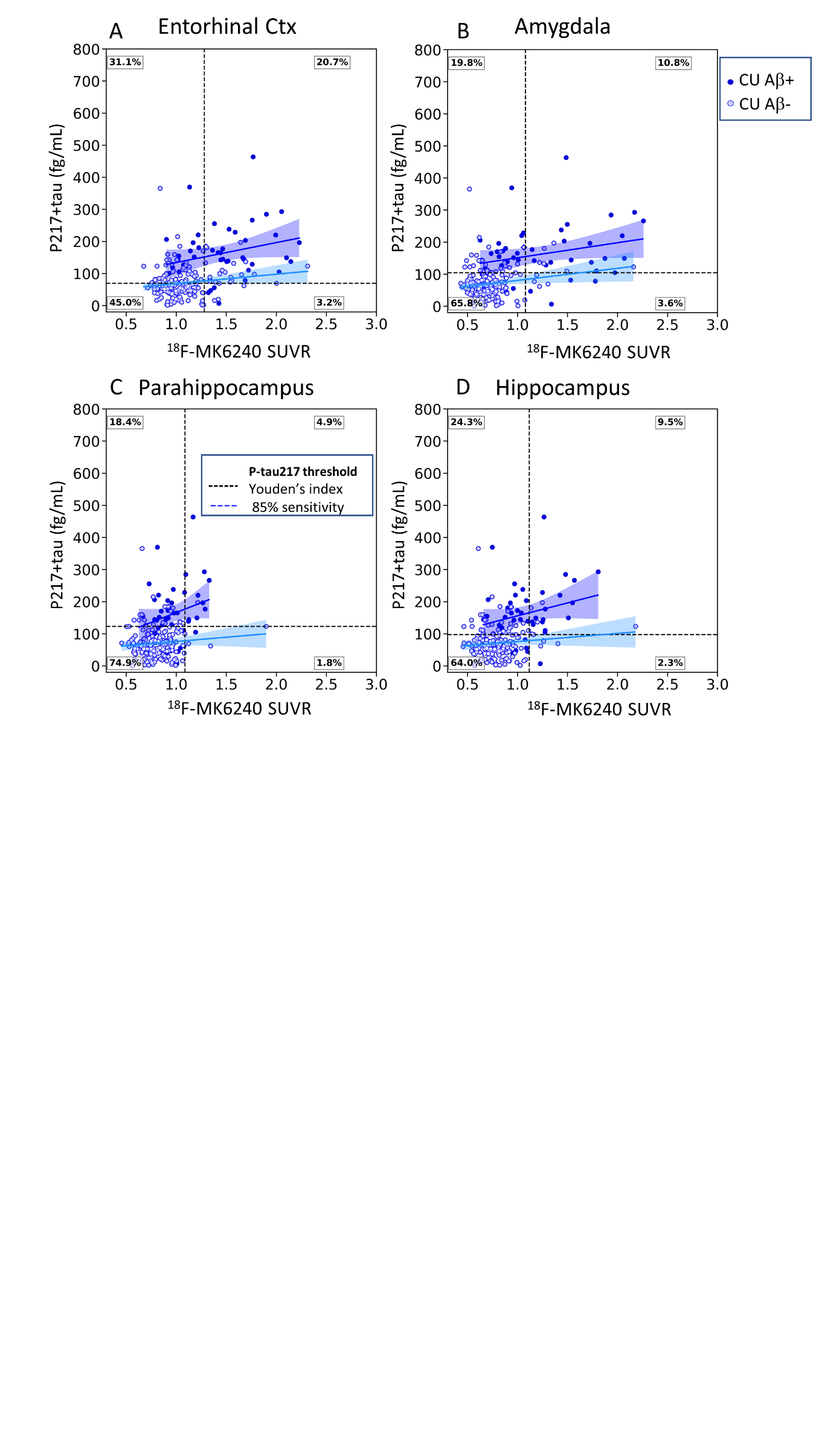

Supplementary Figure 12 Scatter plots of plasma p217+tau concentration versus composite MK6240 SUVR (A: Entorhinal cortex, B) Amygdala, C) Parahippocampus, D) Hippocampus) in cognitively unimpaired individuals, with light colours used to identify A- subjects (CL<25). Thresholds are displayed in fine dashed vertical and horizontal lines. The dashed black vertical line corresponds to the Youden’s index and the blue line the threshold at 85% sensitivity. The diamond shape identified the 2 individuals with an abnormal Aβ- PET scan. The % of individuals in the 4 groups T-p217- T+p217- T-p217+ T+p217+ (Youden) are shown in the outer corners of the plot.

**Z-score analysis**

The P217+tau concentrations were converted into P217+tau z-score, using the CU Aβ- individual as reference group. The statistics reported below were derived from a threshold of 2 standard deviations above the p217+tau mean of the Aβ- CU (164fg/mL).

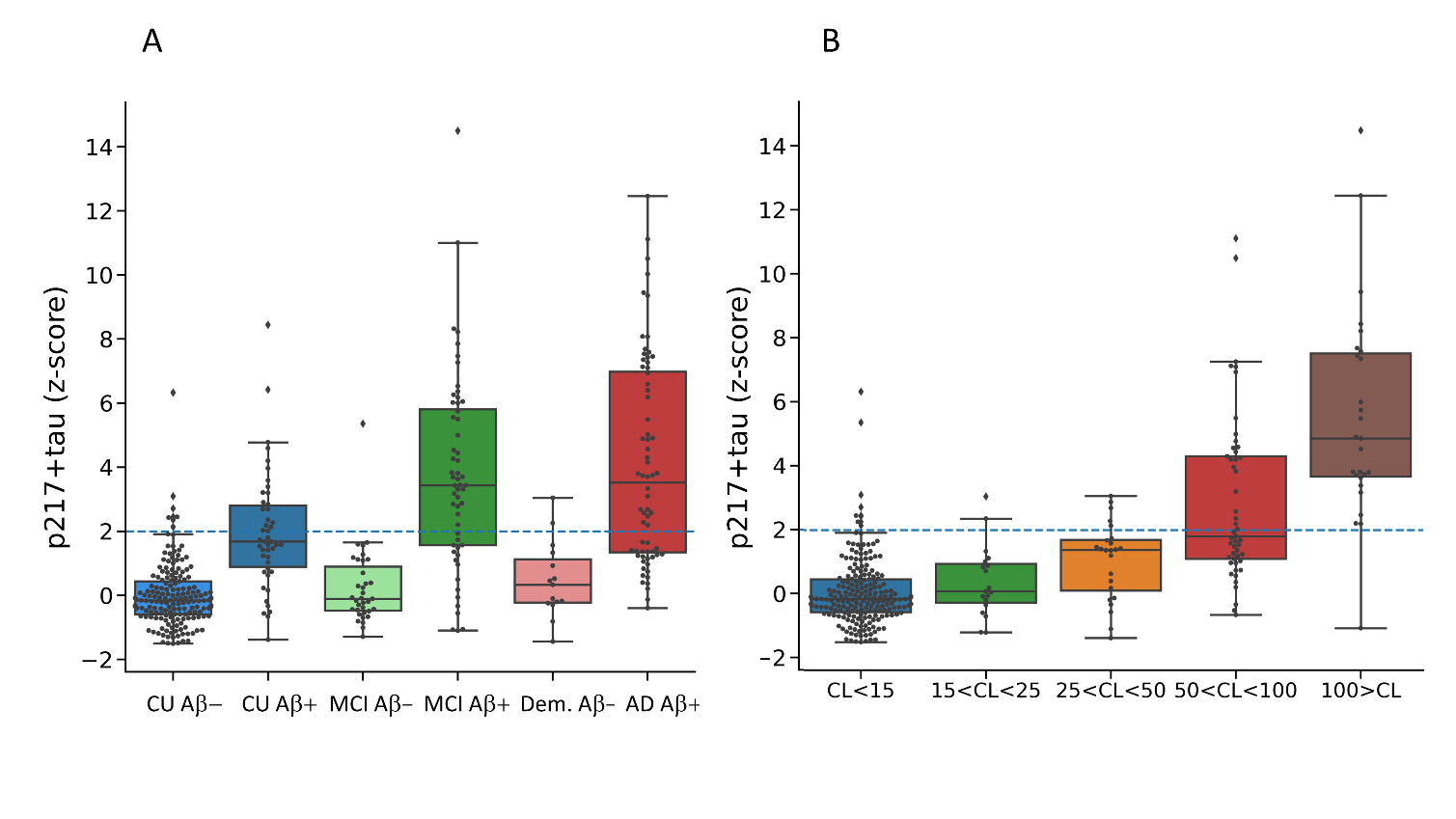

Supplementary Figure 13 Plasma p217+tau concentration z-scores between A) clinical classification and Aβ -PET status and B) levels of Aβ via the CL. The dash blue line corresponds to the two standard deviations from the mean of the Aβ- CU.

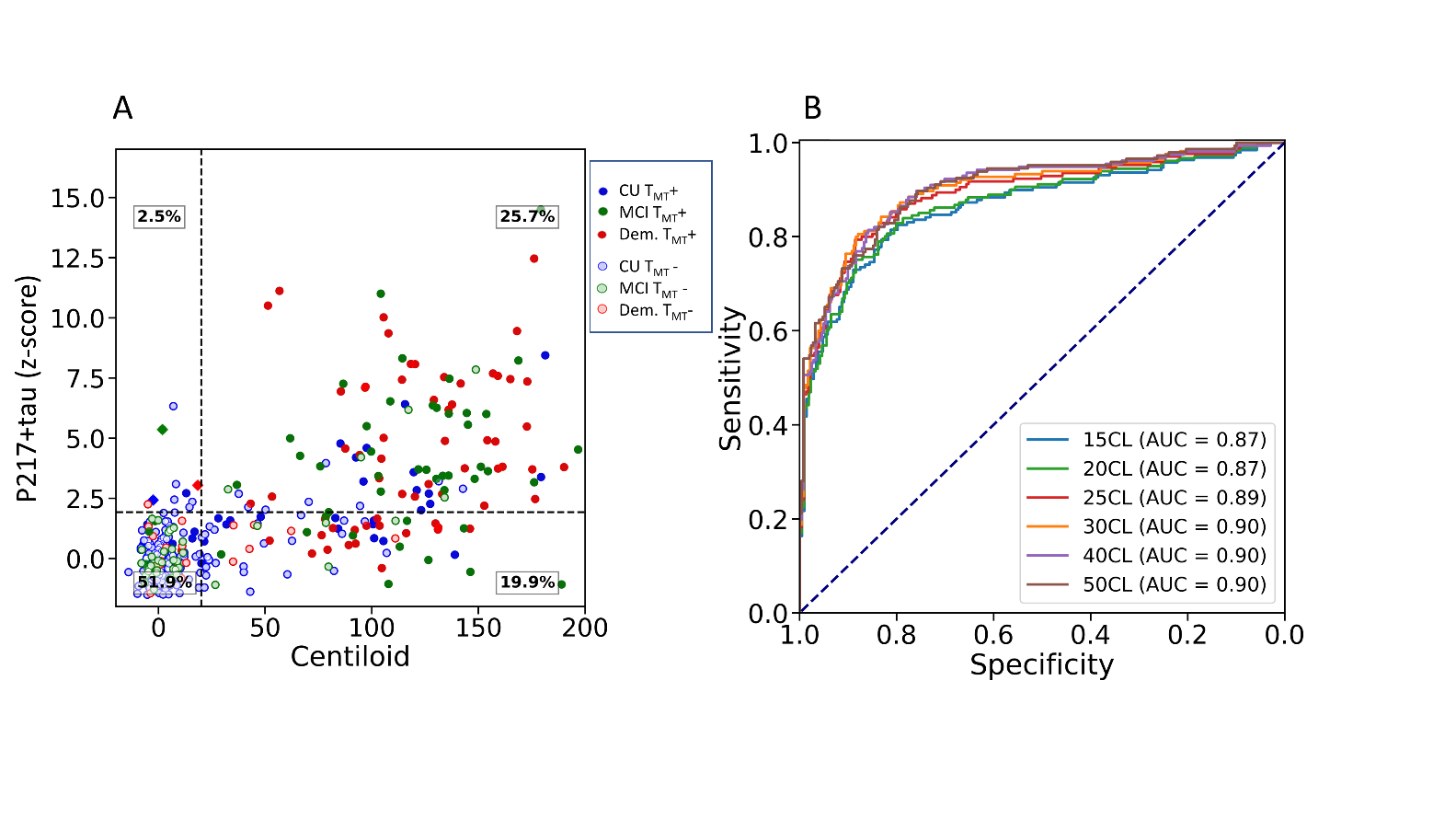

Supplementary Figure 14 A) scatter plot of the p217+tau plasma concentration Z-score versus CL in 6 different colour-coded clinical diagnostic groups, with light colours (opened circles) used to identify T_MT_- subjects. Thresholds are displayed in fine dash vertical and horizontal lines. The horizontal line corresponds to two standard deviations from the mean of the Aβ- CU. The diamond shapes identified the 3 subjects with low Aβ- PET scan and AD-like tau scan. The % of individuals in the 4 groups Aβ-ptau-, Aβ+ptau-, Aβ-ptau+, Aβ+ptau+ are shown in each corner of the plot. B) ROC curves for different thresholds of CL.

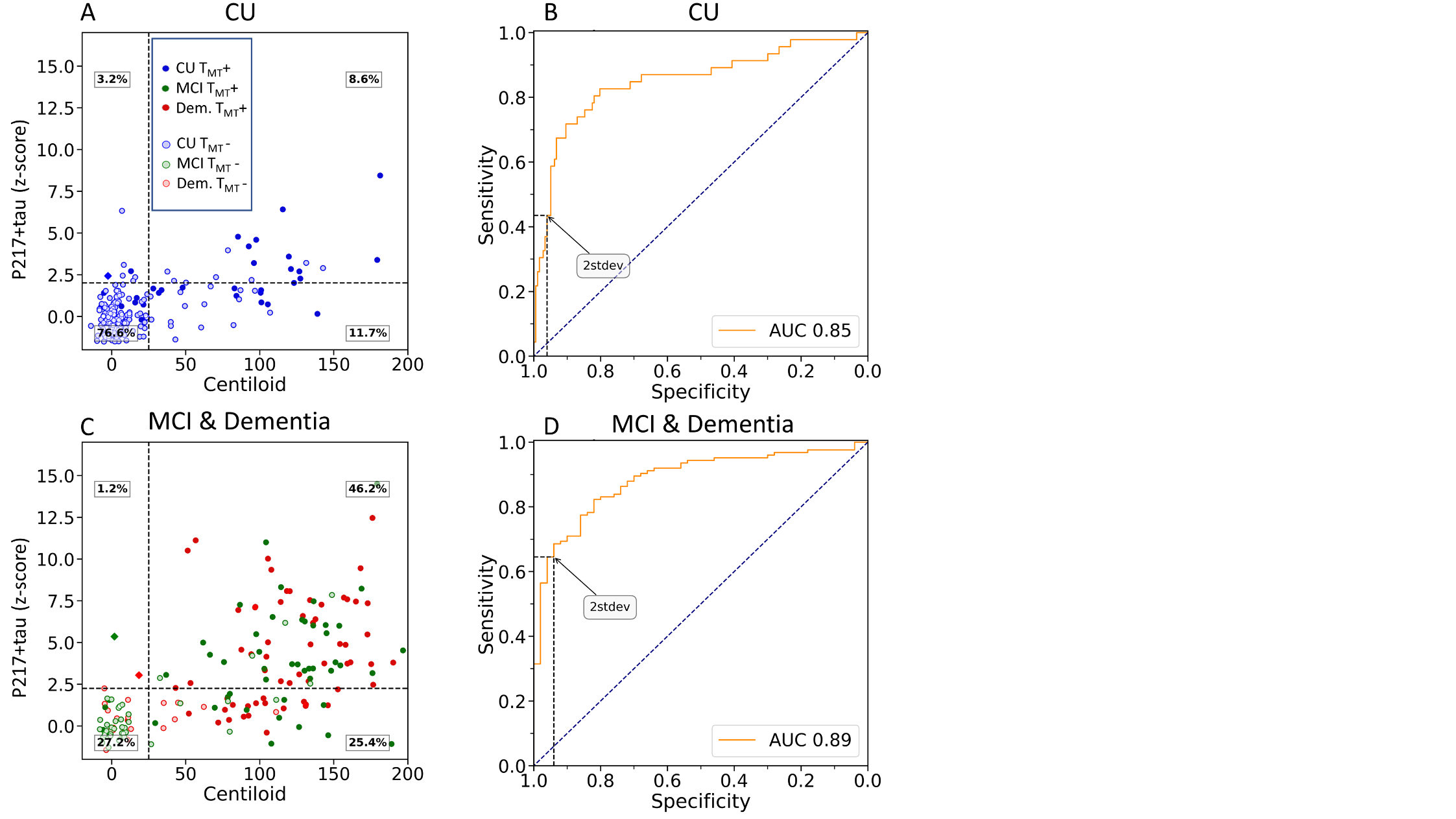

Supplementary Figure 15 A & C) scatter plots of CL versus the plasma p217+tau concentration z-score in 6 different color-coded clinical diagnostic groups (A) CU (blue) and C) CI (green MCI, red dementia)), with light colours used to identify T- subjects. Thresholds are displayed in fine dashed vertical and horizontal lines. The horizontal black line corresponds to two standard deviations from the Aβ- CU individuals. The diamond shapes identified the 3 subjects with a low Aβ- PET scan and AD-like tau scan. The % of individuals in the 4 groups Aβ-p217- Aβ+p217- Aβ-p217+ Aβ+p217+ (Youden) are shown in the far corners of the plot. Figures C & D show the ROC curves for B) CU and D) MCI/AD.

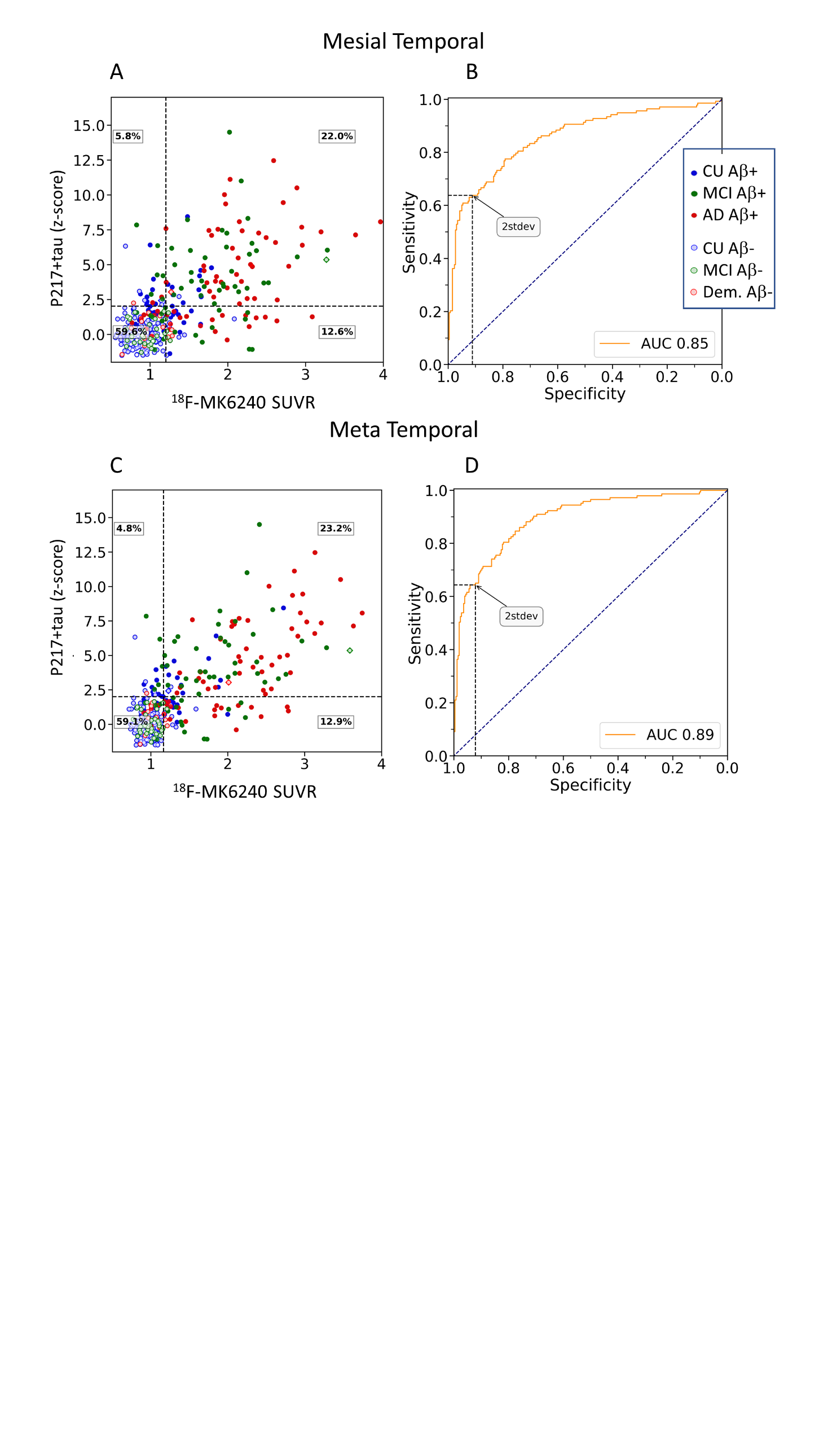

Supplementary Figure 16 A & B, scatter plots of the plasma concentration z-score of p217+tau versus the composite MK6240 SUVR (A) Me, B) Temporal composite) in 6 different color-coded clinical diagnostic groups, with light colours used to identify Aβ- subjects (CL<25). Thresholds are displayed in fine dash vertical and horizontal lines. The horizontal black line corresponds to two standard deviations from the Aβ- CU individuals. The diamond shapes identified the 3 subjects with an abnormal Aβ- PET scan. The % of individuals in the 4 groups T-p217- T+p217- T-p217+ T+p217+ (Youden) are shown in the far corners of the plot. C & D, ROC curve for p217+tau predicting regional PET tau status C) Me & D) Temporal composite.

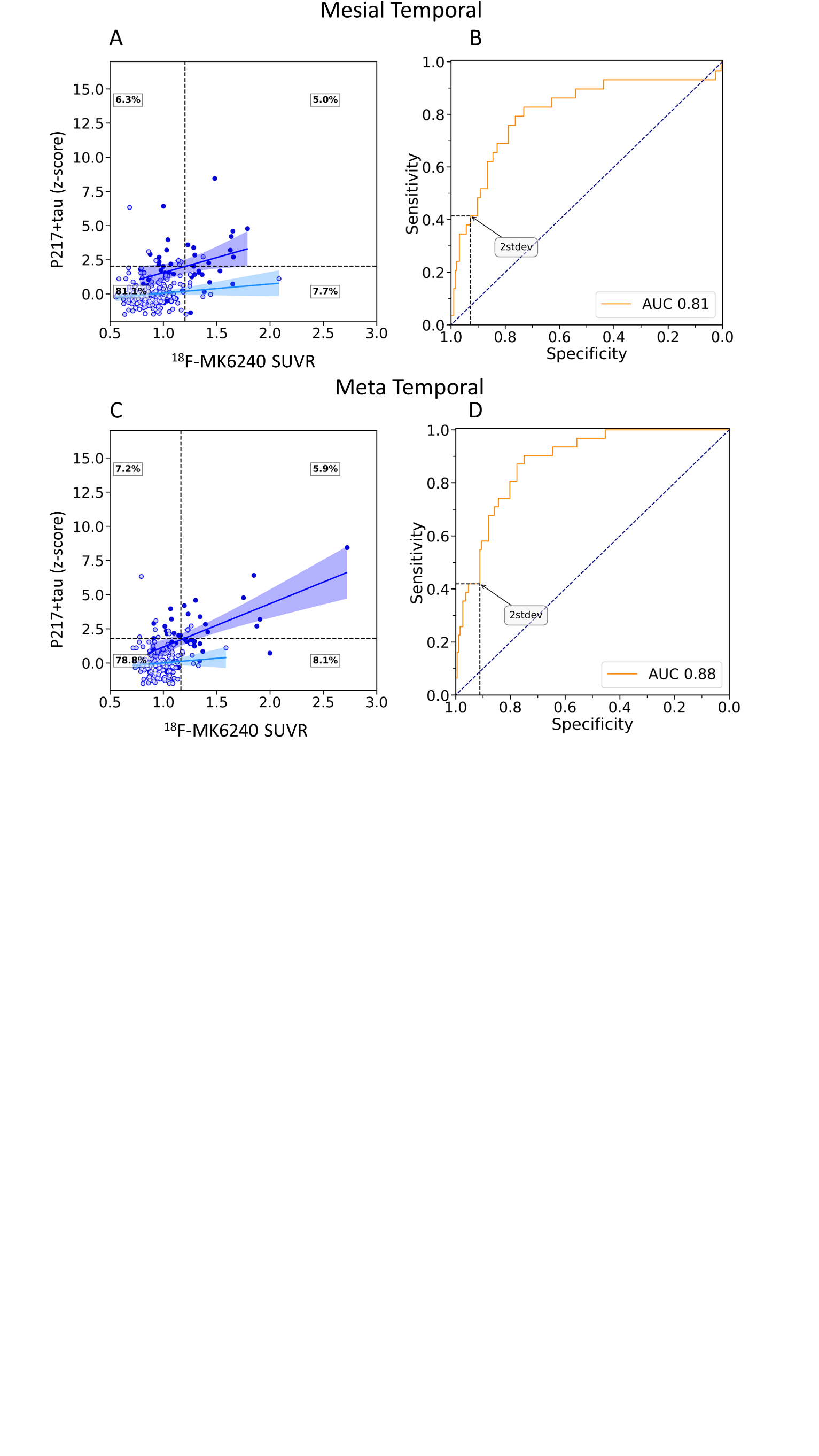

Supplementary Figure 17 A & B: Scatter plots of plasma p217+tau concentration z-score versus composite MK6240 SUVR (A: Me, B: Temporal composite) in cognitively unimpaired individuals, with light colours used to identify Aβ- subjects (CL<25). Thresholds are displayed in fine dash vertical and horizontal lines. The horizontal black line corresponds to two standard deviations from the Aβ- CU individuals. The diamond shape identified the individual with an abnormal Aβ- PET scan. The % of individuals in the 4 groups T-p217- T+p217- T-p217+ T+p217+ (Youden) are shown in the outer corners of the plot. C & D: ROC curve, p217+tau predicting regional PET tau status in CU (C: Me, D: Temporal composite).

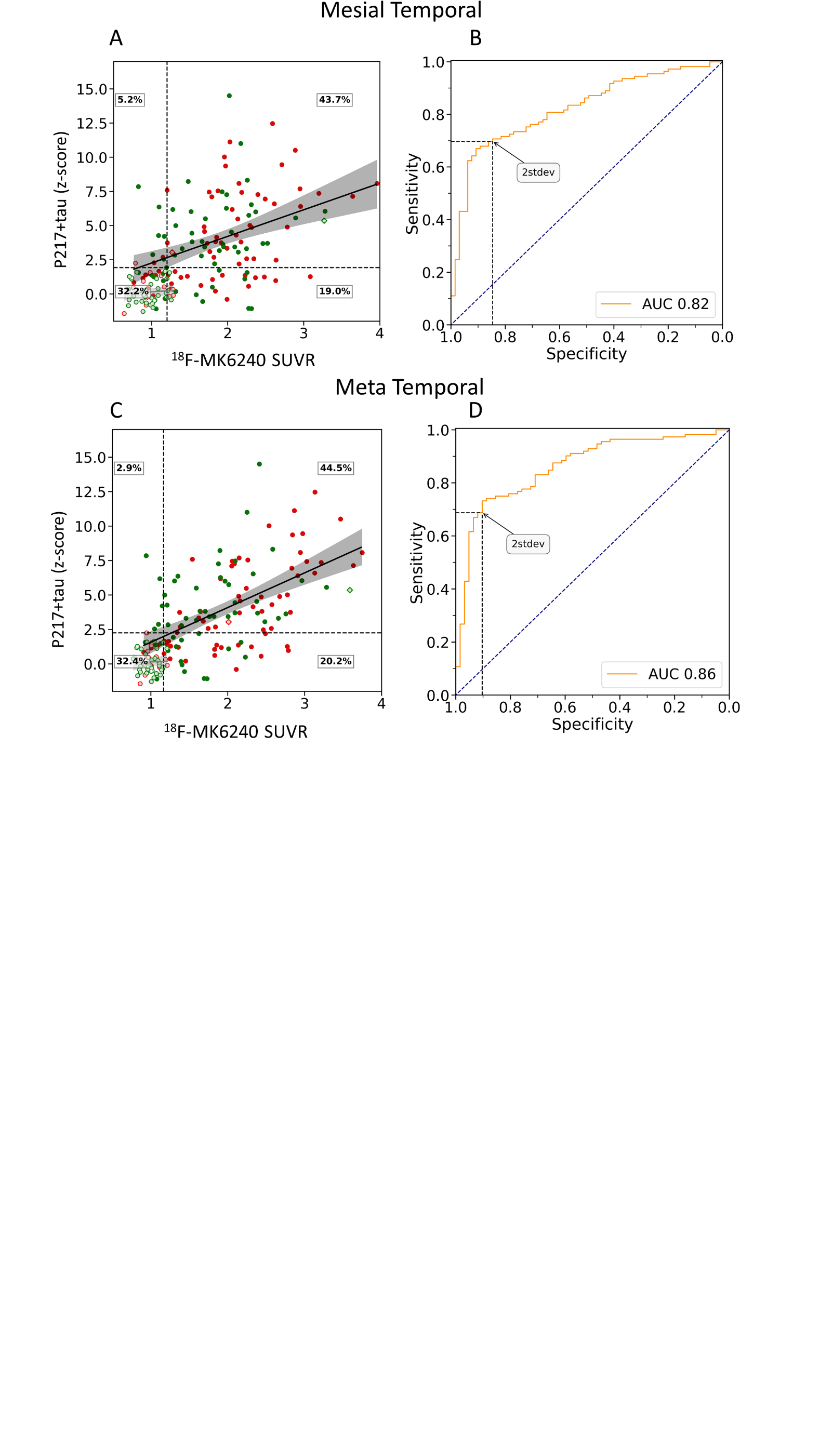

Supplementary Figure 18 A & B: Scatter plots of plasma p217+tau concentration z-score versus composite MK6240 SUVR (A: Me, B: Temporal composite) in cognitively impaired individuals, with light colours used to identify Aβ- subjects (CL<25). Thresholds are displayed in fine dash vertical and horizontal lines. The horizontal black line corresponds to two standard deviations from the Aβ- CU individuals. The diamond shape identified the individual with an abnormal Aβ- PET scan. The % of individuals in the 4 groups T-p217- T+p217- T-p217+ T+p217+ (Youden) are shown in the outer corners of the plot. C & D: ROC curve, p217+tau predicting regional PET tau status in CU (C: Me, D: Temporal composite).

**2 standard deviations threshold**

| **Centiloid** | **AUC**  **(95%CI)** | **Accuracy** | **Sensitivity** | **Specificity** | **PPV** | **NPV** | **Threshold**  **(z-score)** |
| --- | --- | --- | --- | --- | --- | --- | --- |
| **15CL** | 0.87  (0.83-0.90) | 0.77 | 0.55 | 0.96 | 0.93 | 0.7 | 2.0 |
| **20CL** | 0.87  (0.84-0.90) | 0.78 | 0.56 | 0.95 | 0.91 | 0.72 | 2.0 |
| **25CL** | 0.89  (0.86-0.93) | 0.8 | 0.6 | 0.96 | 0.91 | 0.76 | 2.0 |
| **30CL** | 0.9  (0.87-0.93) | 0.82 | 0.62 | 0.96 | 0.91 | 0.78 | 2.0 |
| **40CL** | **0.90**  (0.87-0.93) | 0.82 | 0.63 | 0.95 | 0.88 | 0.8 | 2.0 |
| **50CL** | 0.90  (0.86-0.92) | 0.84 | 0.66 | 0.94 | 0.87 | 0.83 | 2.0 |

Supplementary Table 13: Statistics of the ROC analysis to predict amyloid positivity for different CL threshold of 2stdev.

| **Centiloid** | **AUC**  **(95%CI)** | **Accuracy** | **Sensitivity** | **Specificity** | **PPV** | **NPV** | **Threshold**  **(z-score)** |
| --- | --- | --- | --- | --- | --- | --- | --- |
| **15CL** | 0.78  (0.71-0.86) | 0.78 | 0.33 | 0.96 | 0.78 | 0.78 | 2.0 |
| **20CL** | 0.8  (0.72-0.87) | 0.8 | 0.35 | 0.96 | 0.74 | 0.81 | 2.0 |
| **25CL** | 0.85  (0.78-0.92) | 0.85 | 0.43 | 0.96 | 0.74 | 0.87 | 2.0 |
| **30CL** | 0.85  (0.78-0.93) | 0.87 | 0.47 | 0.96 | 0.74 | 0.88 | 2.0 |
| **40CL** | 0.87  (0.79-0.94) | 0.88 | 0.5 | 0.96 | 0.7 | 0.9 | 2.0 |
| **50CL** | 0.89  (0.82-0.95) | 0.89 | 0.55 | 0.95 | 0.67 | 0.92 | 2.0 |

Supplementary Table 14: Statistics of the ROC analysis to predict amyloid positivity in CU for different CL threshold of 2stdev.

| **Centiloid** | **AUC**  **(95%CI)** | **Accuracy** | **Sensitivity** | **Specificity** | **PPV** | **NPV** | **Threshold**  **(z-score)** |
| --- | --- | --- | --- | --- | --- | --- | --- |
| **15CL** | 0.89  (0.84–0.94) | 0.75 | 0.66 | 0.96 | 0.98 | 0.53 | 2.0 |
| **20CL** | 0.89  (0.83-0.94) | 0.74 | 0.66 | 0.94 | 0.96 | 0.53 | 2.0 |
| **25CL** | 0.89  (0.83-0.94) | 0.74 | 0.66 | 0.94 | 0.96 | 0.53 | 2.0 |
| **30CL** | **0.90**  **(0.85-0.95)** | 0.75 | 0.67 | 0.94 | 0.96 | 0.55 | 2.0 |
| **40CL** | 0.89  (0.84-0.94) | 0.75 | 0.68 | 0.91 | 0.94 | 0.57 | 2.0 |
| **50CL** | 0.88  (0.83-0.93) | 0.77 | 0.7 | 0.9 | 0.93 | 0.62 | 2.0 |

Supplementary Table 15: Statistics of the ROC analysis to predict amyloid positivity in CI for different CL threshold of 2stdev.

| **Region** | **AUC**  **(95%CI)** | **Accuracy** | **Sensitivity** | **Specificity** | **PPV** | **NPV** | **Threshold**  **(z-score)** |
| --- | --- | --- | --- | --- | --- | --- | --- |
| **Mesial Temporal** | 0.85  (0.81-0.88) | 0.8 | 0.61 | 0.91 | 0.79 | 0.8 | 2.0 |
| **Meta Temporal** | 0.89  (0.86-0.92) | 0.82 | 0.64 | 0.92 | 0.82 | 0.82 | 2.0 |
| **Inf. Temporal** | 0.86  (0.83-0.90) | 0.84 | 0.7 | 0.91 | 0.77 | 0.87 | 2.0 |
| **Braak IV** | 0.87  (0.83-0.90) | 0.84 | 0.72 | 0.89 | 0.73 | 0.89 | 2.0 |
| **Braak V** | 0.87  (0.83-0.90) | 0.83 | 0.72 | 0.87 | 0.67 | 0.9 | 2.0 |
| **Braak VI** | 0.82  (0.77-0.86) | 0.8 | 0.7 | 0.82 | 0.48 | 0.92 | 2.0 |

Supplementary Table 16: Results from the ROC analyses to predict tau positivity in separate brain regions at 2 stdev. Positive predictive value (PPV), Negative predictive value (NPV).

| **Region** | **AUC**  **(95%CI)** | **Accuracy** | **Sensitivity** | **Specificity** | **PPV** | **NPV** | **Threshold**  **(z-score)** |
| --- | --- | --- | --- | --- | --- | --- | --- |
| **Mesial Temporal** | 0.8  (0.72-0.87) | 0.85 | 0.39 | 0.92 | 0.44 | 0.9 | 2.0 |
| **Meta Temporal** | 0.88  (0.83-0.92) | 0.86 | 0.42 | 0.93 | 0.48 | 0.91 | 2.0 |
| **Inf. Temporal** | 0.75  (0.65-0.84) | 0.86 | 0.41 | 0.91 | 0.33 | 0.93 | 2.0 |
| **Braak IV** | 0.82  (0.72-0.90) | 0.89 | 0.56 | 0.91 | 0.33 | 0.96 | 2.0 |
| **Braak V** | 0.75  (0.61-0.87) | 0.88 | 0.5 | 0.91 | 0.3 | 0.96 | 2.0 |
| **Braak VI** | 0.66  (0.49-0.81) | 0.87 | 0.36 | 0.89 | 0.15 | 0.96 | 2.0 |

Supplementary Table 17: Results from the ROC analyses to predict tau positivity in CU and in different brain regions at 2stdev. Positive predictive value (PPV), Negative predictive value (NPV).

| **Region** | **AUC**  **(95%CI)** | **Accuracy** | **Sensitivity** | **Specificity** | **PPV** | **NPV** | **Threshold**  **(z-score)** |
| --- | --- | --- | --- | --- | --- | --- | --- |
| **Mesial Temporal** | 0.81  (0.75–0.86) | 0.74 | 0.67 | 0.85 | 0.89 | 0.58 | 2.0 |
| **Meta Temporal** | 0.86  (0.81-0.91) | 0.78 | 0.71 | 0.9 | 0.93 | 0.63 | 2.0 |
| **Inf. Temporal** | 0.86  (0.81-0.9) | 0.82 | 0.76 | 0.89 | 0.91 | 0.73 | 2.0 |
| **Braak IV** | 0.83  (0.78-0.88) | 0.79 | 0.74 | 0.84 | 0.86 | 0.72 | 2.0 |
| **Braak V** | 0.83  (0.78-0.88) | 0.78 | 0.76 | 0.79 | 0.79 | 0.76 | 2.0 |
| **Braak VI** | 0.77  (0.71-0.83) | 0.71 | 0.76 | 0.68 | 0.59 | 0.82 | 2.0 |

Supplementary Table 18: Results from the ROC analyses to predict tau positivity in CI and in different brain regions at 2stdev. Positive predictive value (PPV), Negative predictive value (NPV).
